# Proximity to primary board certification and prior crisis management experience are associated with anesthesiologists’ performance in high-fidelity acute care scenarios

**DOI:** 10.64898/2026.09.10.26362280

**Authors:** Matthew B. Weinger, William R. McIvor, Jason M. Slagle, Josh DeClercq, Caoimhe C. Duffy, Jeffrey Huang, Matthew S. Shotwell

## Abstract

**Introduction:** Consistent high-level performance during perioperative critical events is essential for achieving optimal patient outcomes. We sought to better understand factors associated with anesthesiologists’ performance during simulated crisis management.

**Methods:** Sixty clinically active board-certified anesthesiologists (BCAs), 22 junior (6 to 18 mo. training) and 19 senior (24 to 48 mo.) anesthesia trainees participated in standardized, high-fidelity simulated out-of-OR crisis events. Participants completed demographic and practice attributes surveys, and then performed four different 15-minute scenarios. Trained, board-certified raters scored randomly assigned video-recorded performances using checklists and holistic scaled ratings. A principal component analysis summarized survey responses related to participants’ clinical, simulation, and crisis management experience. Mixed-effects regression was used to examine the relationship of performance ratings to the summarized variables, adjusting for participant attributes, study site, and scenario.

**Results:** The participants were 55% male, 38±8 years old and had 8.8±6.7 years of experience (training included). The BCAs were 67% community practitioners. Generally, senior trainees received the highest scores while junior trainees received the lowest. There was a strong association between overall performance and combined years of training and experience (p = 0.008). Experience with prior crises (p=0.040), but not clinical practice attributes (p=0.97) or prior simulation experience (p=0.94), was associated with higher rated performance. High-performing participants were more likely to call for assistance, physically examine the patient, order additional diagnostic tests, escalate therapy (e.g., greater hemodynamic support or antibiotic coverage), broaden the differential, discuss concerns with a colleague, and provide both temporizing and definitive treatment.

**Discussion:** This simulation-based study found differences in performance ratings on perioperative crisis events based on experience and prior exposure to challenging clinical situations. These findings corroborate prior work, validate the use of simulation for assessment, and reinforce the value of ongoing crisis management exposure and training to improve patient care.

## Introduction

Healthcare professionals are expected to competently manage acute, including rare, critical events; yet deficiencies occur during actual events.^1,2^ Managing critical events such as cardiorespiratory arrest or anaphylactic shock demands both technical (e.g., correct diagnosis and therapy) and nontechnical (e.g., teamwork/communication) skills. In these high-stakes situations, anesthesiologists must apply Crisis Resource Management (CRM) principles^3,4^ while simultaneously stabilizing the patient through iterative diagnose-and-treat cycles.^5,6^ Skill deficiencies can lead to “failure-to-rescue” and increased risk of adverse outcomes.^1,2,7,8^

Anesthesiologists’ performance varies during critical events, and previous high-fidelity simulation studies show as many as 30% manage these situations suboptimally.^9,10^ Conversely, the majority perform well, suggesting that antecedent factors, such as specific training and clinical experience, may influence performance. Although physician skill decay over time,^11–13^ including of crisis management,^9,10,14^ has been well described, it is not routinely assessed after board certification, and the factors that mitigate decay in experienced anesthesiologists remain unclear.

High-fidelity simulation is a practical method to identify anesthesiologists likely to struggle during a critical event. Prior literature highlights simulation’s value in assessing and providing formative feedback on technical and nontechnical skills in healthcare through objective, standardized, and replicable crisis scenarios.^4,15–18^ High-fidelity simulation provides a standardized platform to ensure fair and reliable performance assessment.^19^ However, it is unclear how clinicians’ demographics, clinical practice attributes, prior exposure to clinical crises, and previous simulation experience correlate with individual’s performance in simulated crisis scenarios.

In a prior multisite study,^9^ board-certified anesthesiologists (BCAs) were assessed in standardized critical intraoperative scenarios during maintenance of certification in anesthesiology (MOCA™) courses. Almost one-third of the study encounters were rated as “poor” overall holistic technical and nontechnical performance. Moreover<u>, specific patterns</u> <u>of technical and nontechnical performance deficiencies (i.e. ‘gaps’) were identified</u>.^9,20,21^ However, t<u>his “MOCA</u> Simulation Study” (MOCA-Sim) was not designed to assess individual overall performance nor to delineate the factors associated with low versus high-performance scores.

We undertook a new study of BCAs and resident anesthesiologists performing in simulated scenarios to test the *a priori* hypotheses that: **Hypothesis 1**. Total years of clinical anesthesiology training and experience will be positively correlated with performance in simulated crises;^9^ **Hypothesis 2.** Prior crisis management (in actual or simulated experiences) will be positively correlated with performance in simulated crises; and **Hypothesis 3.** Previously identified clinical performance gaps^9,20^ will be replicated, with the potential of identifying new gaps. The study was also designed to explore decision-making strategies through cognitive interviews that followed each scenario – findings to be reported in a subsequent publication.

## Methods

This study was approved through a SingleSite IRB (#181006) with Vanderbilt University serving as the Coordinating Center.

### Study sites

This prospective observational multisite cohort study was conducted at four high-fidelity simulation centers – Vanderbilt University Medical Center’s Center for Experiential Learning and Assessment (CELA at VUMC, Nashville, TN), The Winter Institute for Simulation Education and Research (WISER) at the University of Pittsburgh Medical Center (UPMC, Pittsburgh, PA), UNM BATCAVE Simulation Education and Training Center at the University of New Mexico Health Sciences Center (UNM, Albuquerque, NM), and the Clinical Simulation Center (CSC) at the Pennsylvania State College of Medicine (PSU, Hershey, PA).

### Participants

We recruited BCA participants through state and local specialty societies, practice groups, and the ‘Physician Anesthesiologist Mom’ Facebook group. Trainee participants came from residency and fellowship programs at or near our study sites. A total of 102 of 104 enrolled anesthesiologists (61 BCAs and 41 residents) participated in the study between December 2020 and January 2022 (see **SDC-1. Figure S1**). Participants provided written informed consent. Participants were paid and BCAs received 7.5 hours of ASA Patient Safety CME and MOCA Part IV credit.

### Inclusion/Exclusion criteria

BCA participants were clinically active (≥50%) in any anesthesia subspecialty (excluding chronic pain) and had taken their primary certification exams between 2002 and 2021. Those in ‘academic practice’ self-reported consistent supervision of residents. BCAs Anesthesia residents who had ≥6 months of clinical anesthesia training and had not taken the CA2 In-Training Examination offered by the American Board of Anesthesiology (ABA, Raleigh NC) were considered “Junior” trainees. Those who had ≥2.5 years of clinical anesthesia training but were not yet board-certified, were considered “Senior” trainees.

### Demographic survey (see **SDC-2**)

We designed and pilot tested an expanded version of our previous anesthesiologist professional attribute survey^9^ to include additional training, experience, and practice attributes that might affect crisis management performance. The final survey contained seven modules – 1) Basic Demographics, 2) Prior Clinical Training, 3) Current Practice Attributes, 4) Exposure to Challenging Clinical Situations, 5) Prior Simulation-based Training, 6) Hazardous Attitudes,^22^, and 7) the Maslach Burnout Inventory.^23^

### Scenario development and standardization

#### Scenario selection

Four 15-minute high-fidelity simulation scenarios (**Table 1**) were designed and pilot tested to: 1) be challenging but manageable by BCAs; 2) require significant decision making and teamwork skills; 3) occur in pre- or post-operative, non-operating room, settings; 4) have several plausible diagnoses; 5) be performed individually, with help available from standardized embedded participants (SEPs) portraying clinical team members according to strict behavioral scripts; and 6) end with a hand-over of care to another physician (via phone) to capture participants’ end-of-scenario thought processes. Scenario content was vetted by an independent multidisciplinary perioperative Subject Matter Experts panel (SMEs, **SDC-3. Table S1**), who also delineated the expected differential diagnoses and the scoring criteria (see below). Henceforth, scenario names will be *italicized*.

**Table 1.**
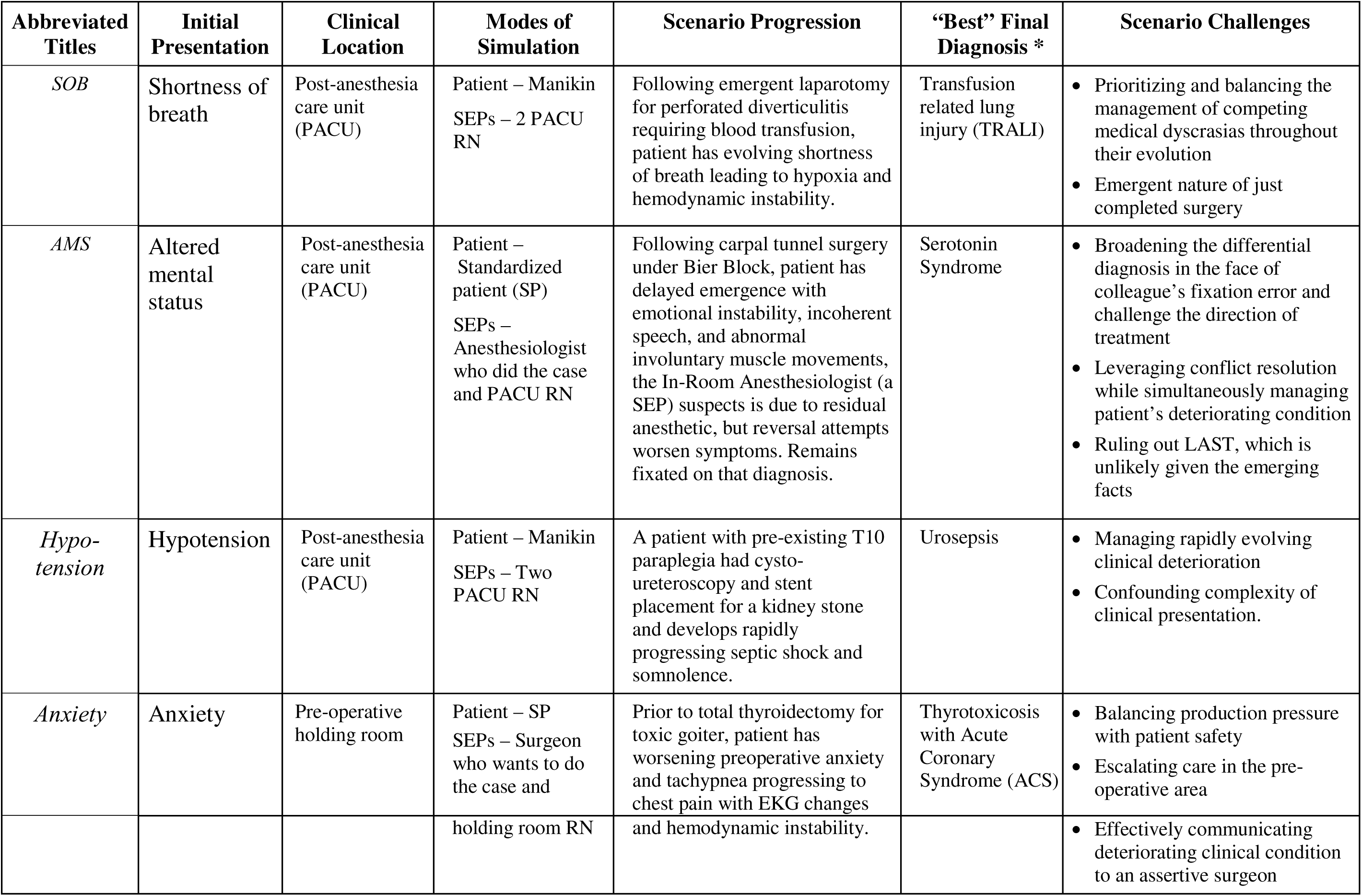
Scenario Descriptions.

| Abbreviated Titles | Initial Presentation | Clinical Location | Modes of Simulation | Scenario Progression | “Best” Final Diagnosis * | Scenario Challenges |
| --- | --- | --- | --- | --- | --- | --- |
| *SOB* | Shortness of breath | Post-anesthesia care unit (PACU) | Patient – Manikin<br>SEPs – 2 PACU RN | Following emergent laparotomy for perforated diverticulitis requiring blood transfusion, patient has evolving shortness of breath leading to hypoxia and hemodynamic instability. | Transfusion related lung injury (TRALI) | • Prioritizing and balancing the management of competing medical dyscrasias throughout their evolution<br>• Emergent nature of just completed surgery |
| *AMS* | Altered mental status | Post-anesthesia care unit (PACU) | Patient – Standardized patient (SP)<br>SEPs – Anesthesiologist who did the case and PACU RN | Following carpal tunnel surgery under Bier Block, patient has delayed emergence with emotional instability, incoherent speech, and abnormal involuntary muscle movements, the In-Room Anesthesiologist (a SEP) suspects is due to residual anesthetic, but reversal attempts worsen symptoms. Remains fixated on that diagnosis. | Serotonin Syndrome | • Broadening the differential diagnosis in the face of colleague’s fixation error and challenge the direction of treatment<br>• Leveraging conflict resolution while simultaneously managing patient’s deteriorating condition<br>• Ruling out LAST, which is unlikely given the emerging facts |
| *Hypotension* | Hypotension | Post-anesthesia care unit (PACU) | Patient – Manikin<br>SEPs – Two PACU RN | A patient with pre-existing T10 paraplegia had cysto-ureteroscopy and stent placement for a kidney stone and develops rapidly progressing septic shock and somnolence. | Urosepsis | • Managing rapidly evolving clinical deterioration<br>• Confounding complexity of clinical presentation. |
| *Anxiety* | Anxiety | Pre-operative holding room | Patient – SP<br>SEPs – Surgeon who wants to do the case and | Prior to total thyroidectomy for toxic goiter, patient has worsening preoperative anxiety and tachypnea progressing to chest pain with EKG changes | Thyrotoxicosis with Acute Coronary Syndrome (ACS) | • Balancing production pressure with patient safety<br>• Escalating care in the pre-operative area |
|  |  |  | holding room RN | and hemodynamic instability. |  | • Effectively communicating deteriorating clinical condition to an assertive surgeon |

#### Standardized Embedded Participants

Scripts were created for SEPs that defined their roles, including gender, age range, appearance, information of which they were aware, and motivations (e.g., ‘you feel rushed as the surgeon is pushing to get their case started’). Standardized descriptions of the patient or situation were created and responses to all likely participant inquiries were scripted (e.g., “do not say ‘sweaty’, say ‘appears in distress’”).

#### Scenario attributes and events

Vital sign changes, responses to interventions, patient states, and SEP’s responses to questions or key observations were scripted. ‘Bail outs’ to help participants who were not progressing during the scenarios were scripted and standardized to ensure everyone saw and managed all essential scenario phases. Scenario-specific initial settings, entry conditions, and time zero were established. Scenarios ended with each participant being prompted to call another clinician to transfer the patient’s care. Standardized clinical documents provided essential patient information and diagnostic test results. Rules for scenario operators provided standardized experimental conditions (**SDC-4**). Vital sign changes and responses to interventions were scripted and, to the extent possible, automated to optimize standardized scenario delivery.

#### Pilot testing and “Sign off”

All tools, processes, and scenarios were extensively pilot tested at different sites. Prior to study enrollment at each site, investigators, including the scenario authors, reviewed a recorded pilot run of each scenario to calibrate delivery accuracy.

### Study day (Figure 1)

At the start of the study day, the previously emailed REDCap™-based eConsent and e-Survey responses were reviewed and confirmed. Participants were oriented regarding the study day, rules, simulated clinical settings, and the cognitive interview process^24^. Participants were told that the scenarios were based on actual cases, were not intended to be deceptive, and that COVID was not in the differential diagnoses. They were instructed to react as they would in their real-world practice and received structured training on how to interact with manikins and standardized patients. Finally, participants performed a simple non-study scenario, using study forms and processes, to test and receive feedback on their understanding of the study’s rules and methods.

**Figure 1.**
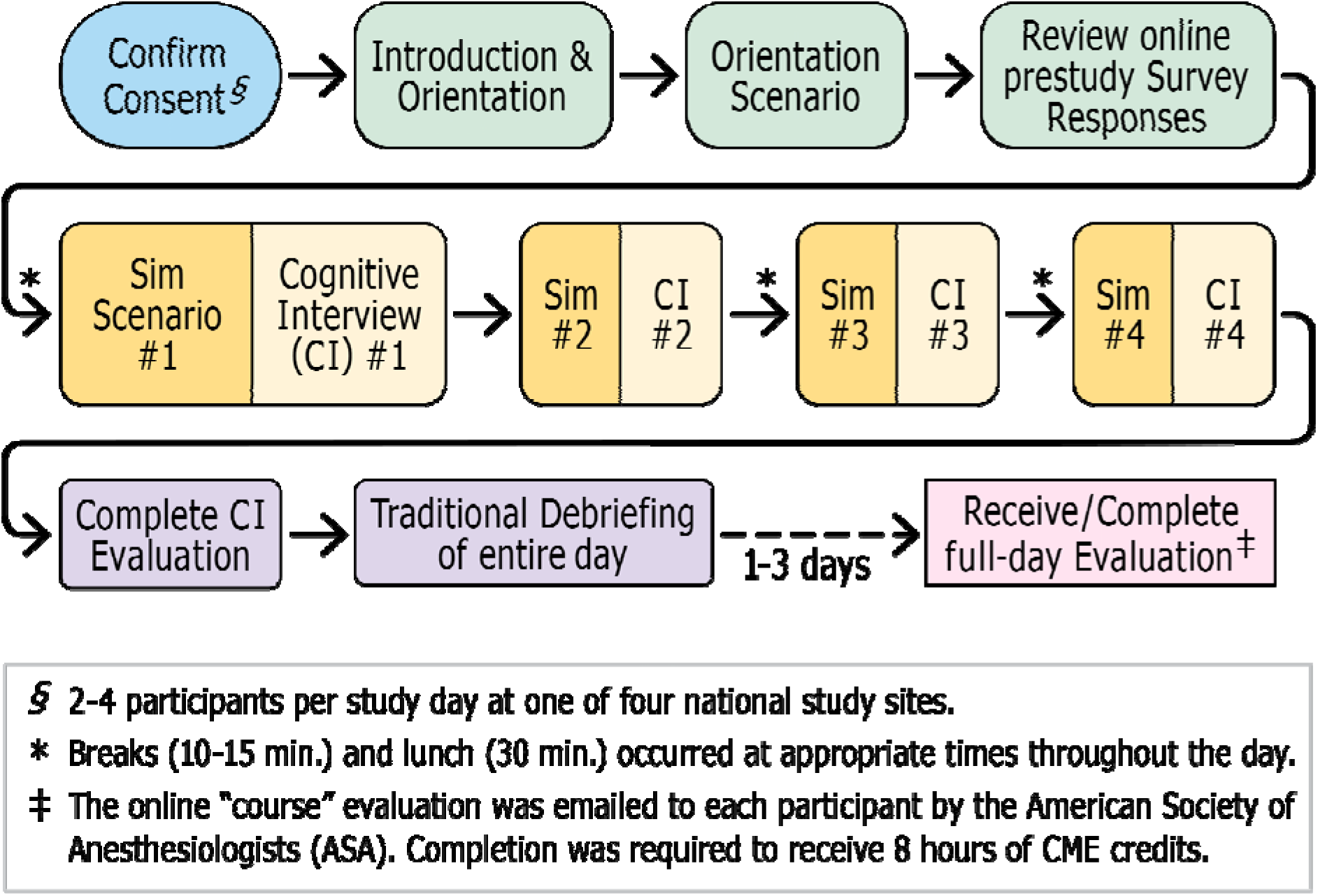
Study Day Flow. Each participant completed an and the comprehensive demographic/practice survey before coming. Upon arrival to the 8-hour study session, the previously completed online written informed consent was confirmed. The participant was then oriented to the study, the day’s activities, and the simulation environment. They performed in a realistic orientation scenario (often with other study participants). Each participant then did 4 sequential 15-to 20-minute study scenarios, each one followed by a 40-minute cognitive interview. Upon completion of the fourth simulation-interview pair, the participant completed survey’s and received an educational debriefing with at least one expert anesthesiology faculty.

#### Scenario order and delivery

Each participant completed all four simulation scenarios, which were each followed immediately by a 40-minute cognitive interview (CI),^24^ whose findings will be presented separately. Scenario order was randomized with blocked allocation at two study sites based on scenario-logistical constraints. After finishing the study, participants completed an eSurvey about their CI experience, participated in an educational debriefing session, and provided an ‘course evaluation’ of their experience (**SDC-5**).

### Videorecording and processing

All encounters were video recorded using two orthogonal camera views plus a vital-signs monitor feed. These were uploaded into VidARA™, a custom video data management software application (**SDC-6**) and automatically reformatted into a quad-picture view. Participants’ VidARA™ videos and subsequent rating data (see below) were linked to their REDCap™ survey data.

### Performance measures

*Clinical Performance Elements (CPEs)* are observable, scenario-specific checklist items (scored ‘present or absent’)^9,20^ based on behaviors expected of excellent participants. Using a modified Delphi process, the SMEs iteratively developed and reached consensus on the CPEs for each scenario (**SDC-7** and **Table 3**).

*Behaviorally anchored ratings scales (BARS)*^25^ were modified from our prior work^9^ to better align with this study scenarios’ performance expectations. Five domains were assessed: Vigilant Observation, Situation Awareness and Re-evaluation, Diagnostic Processes, Action Processes, and Communication (**SDC-8**). Each BARS had an extensive set of observable behaviors for different performance levels in that domain. BARS were scored on a 9-point scale from 1 (worst) to 9 (best).^9^

#### Global ratings

Raters used 9-point scales to assess technical (medical), nontechnical (behavioral), and overall performance, with the ranges of low (1-3), medium (4-6) and high (7-9).^9^ Finally, a binary rating, ‘Did this participant perform at the level of a consultant anesthesiologist?’ was made.

### Video ratings

#### Raters

After a national call for rater nominations, 10 individuals met our *a priori* criteria: 1) ABA certification with ≥3 years of post-certification clinical experience; 2) Prior experience in and understanding of simulation methods; and 3) Substantial experience providing education, training, and/or assessment of trainees and practitioners.

#### Rater training and certification

Raters were trained and certified as previously described.^9^ BCA study team members, overseen by the rater trainers, established consensus ratings on 38 study videos representing the full range of performances in each scenario. Raters participated in two-hour virtual training orientation sessions and received individualized training and targeted feedback for each scenario. Raters were certified when their CPE ratings matched the established consensus ratings, their BARS scores were within one point, and their holistic ratings fell within the same initial range (low, medium, high). Five individuals were certified to rate at least two different scenarios.

#### Rating process

Raters received randomized batches of video recordings. Raters did not score a performance if they recognized a participant. A random 20% of videos were assigned to two different raters. Raters viewed the performances in a single pass, creating a time-stamped log of annotations and CPEs performed. CPEs were scored as present if the participant, or a SEP under their direction, performed it anytime during the encounter. Then, raters scored the five BARS domains and made holistic performance assessments. Raters were compensated for each encounter successfully completed.

### Statistical analysis and modeling

#### Rater reliability

Intra-rater reliability was evaluated using the intraclass correlation coefficient (ICC) for scaled scores while Cohen’s kappa was used for binary measures. Inter-rater reliability was assessed using Krippendorff’s alpha to accommodate different numbers of raters across different measurement levels. An ordinal specification was applied for the different scaled performance scores. A nominal specification was used for the binary consultant level performance (CLP) and CPEs. Criteria for Cohen’s kappa, ICC, and Krippendorff’s alpha are found in **SDC-10**.

#### Covariates

To reduce the >100 experience-related variables, while retaining the most relevant features, we applied principal component analysis (PCA) to pre-selected variables (see **SDC-9** for details). PCA was conducted separately to derive participants’ composite measures in: 1) clinical practice; 2) crisis management; and 3) simulation experience. For each domain, the first two principal components were retained for subsequent modeling. This reduced the experience covariates to six principal component scores (two per domain).

#### Performance score reporting and modeling

Continuous outcomes and participant attributes were summarized using mean and standard deviation (SD) or median and interquartile range (IQR). Categorical variables were summarized with frequencies and percentages. The significance of unadjusted associations was quantified using the Wilcoxon-Kruskal-Wallis test for continuous variables and the Pearson chi-square test or Fisher’s exact test for categorical variables.

Linear mixed-effects regression was used to quantify the associations separately between technical, non-technical, and overall performance ratings (dependent variables) and the composite principal component variables for clinical, crisis management, and simulation experience (independent variables), adjusting for experience cohort (Junior, Senior, BCA), gender, and simulation scenario. A random intercept indexed by participant and study site accounted for heterogeneity in performance among participants. Quantitative variables were assumed to have a linear effect. We tested the null hypothesis that none of the composite variables representing experience affected performance scores, using a two degree-of-freedom Wald (‘chunk’) test. The significance of each individual independent variable was also assessed using Wald tests.

A generalized linear mixed-effects model (GLMM) with a logit link function was used to model consultant level performance, adjusting for the same covariates. Mixed-effects models used all available data assuming that data were missing at random, so that the two participants with isolated missing assessments were included.

We also modeled the number of years of training and experience as a continuous variable. The above analyses were repeated, hypothesizing a nonlinear association between the outcome and continuous experience. Nonlinearity was modeled using a restricted cubic spline with four knots, and overall tests of association were conducted using Wald tests. Estimated marginal effects were generated across the range of years of training/experience and visualized using predicted effects plots with corresponding confidence intervals.

#### Clinical Performance Elements (CPEs)

To account for different number of CPE items per scenario, we used a binomial generalized linear mixed-effects model with a logit link, where the number of correct responses was modeled as a binomial random variable conditional on the total number of items per scenario. Scenario was included as a fixed effect to estimate differences across the prespecified scenarios, and participant was included as a random intercept to account for within-subject correlation. The association between overall performance ratings and proportion of CPEs present was assessed using a linear mixed effects model, with participant ID and scenario included as random intercepts.

## Results

### Participant demographics and attributes

Among 102 anesthesiologists who participated in the study (**SDC-1. Figure S1**), 56 (55%) were male and 78 identified as White (**Table 2**). Median age was 36 (IQR 32 – 43), with a median 7.3 (IQR 2.8 – 13.3) years of residency training and relevant work experience. There were 22 Juniors, 19 Seniors, and 61 BCAs (41 Community and 20 Academic). Thirty BCAs had previously completed a MOCA Simulation Course at an ASA endorsed center (49% of BCAs and 29% of all participants).

**Table 2.** Participant Demographics by Experience Group.

| Characteristic | N | BCA<br>(n=61) | All US<br>BCAs<br>(n=22,309)<sup>†</sup> | Senior<br>(n=19) | Junior<br>(n=22) | All US<br>Residents<br>(n=6371) ‡ | Overall<br>(n=102) |
| --- | --- | --- | --- | --- | --- | --- | --- |
| Sex, female | 102 | 32<br>(52.5%) | 33.8% * | 6<br>(31.6%) | 8<br>(36.4%) | 33.4% § | 46<br>(45.1%) |
| Age (years) | 102 | 42.3±7.6<br>(42, 36-48) | 42.7±5.9<br>(42, 30-76) | 32.2±3.2<br>(32, 30-32) | 31.4±4.1<br>(30, 27-42) |  | 38.1±8.2<br>(36, 32-43) |
| Race, non-white | 100 | 13<br>(21.7%) | 32.5% | 5 (27.8%) | 4 (18.2%) | 43.5% § | 22 (22%) |
| Experience<br>(including training)<br>(yrs) | 102 | 13±5.5<br>(12.1, 8.2-17) | 13.5±4.1<br>(13.2, 4.2-21.5) | 3.6±0.9<br>(3.5, 3.0-4.0) | 1.6±0.5<br>(1.4, 1.2-2.2) |  | 8.8±6.7<br>(7.3, 2.8-13.3) |
| Community (%) |  | 41<br>(67.2%) |  |  |  |  |  |
| Total hrs/wk<br>worked | 102 | 51.0±12.4<br>(50, 45-60) |  | 59.1±8.3<br>(60, 55-65) | 61.4±7.0<br>(62.4, 60-65) |  | 54.8±11.6<br>(55, 50-60) |
| Night calls/mo | 93 | 2.3±1.9<br>(2, 0-4) |  | 4.1±2.2<br>(4, 2-5) | 2.9±1.7<br>(3, 2-4) |  | 2.8±2.0<br>(3, 1-4) |
| Burnout? | 102 | 40<br>(65.6%) |  | 11<br>(57.9%) | 17<br>(77.3%) |  | 68<br>(66.7%) |

Continuous values reported as mean (SD) and median (IQR)
<sup>†</sup> = Data from ABA website for 2022 (see <http://>). Blank cells indicates data not available.
‡ = Data from ACGME Report on Resident (see https://www.aamc.org/data-reports/students-residents/data/report-residents/2021/executive-summary). Blank cells indicates data not available.
\* = Significant difference in study population vs. national BCA population (p=0.002).
NB: Difference in race, p=0.053
§ = Significant difference in study population vs. national resident population (p=0.0XX).

### Experience covariates

As described in detail in **SDC-9**, the first two principal components (PCs) explained 43 – 46% of the variance in each of the three experience covariates. For *Crisis Management* (7 variables), PC1 primarily reflected recent exposure to challenging clinical situations and work involving emergent or unstable patients. PC2 largely represented prior formal training in crisis management. For *Simulation Experience* (7 variables), PC1 captured participants’ prior involvement in crisis simulation activities, either as participants or instructors. PC2 was characterized mainly by experience in MOCA simulation or the ASA’s SimStat™ screen-based trainer. For *Clinical Experience* (10 variables), PC1 was associated with work in tertiary care medical centers, supervision of trainees, and performing one’s own cases. PC2 reflected the breadth of subspecialties practiced and weekly hours of clinical work.

### Rater reliability

Two videos could not be rated due to technical difficulties (one *SOB* and one *Anxiety*). Two *AMS* encounters were excluded from the intra-rater reliability (IRR) analysis due to obvious discrepancies. Overall, IRR was good to excellent (**SDC-10. Table S4**), being overall above 0.74 across the scoring items. By scenario, CPEs had the highest IRR (range 0.82 to 0.93), while non-technical ratings had the lowest (0.48 to 0.90). Inter-rater reliability was overall a bit lower ranging from 0.54 for BARS to 0.85 for CPEs.

### Performance score distributions

Across all encounters, a median of 57% (interquartile range [IQR], 52 to 63%) of the CPEs were observed. The highest frequency of observed CPEs was in the *AMS* (67% [IQR, 56 to 78%]) and lowest in the *SOB* scenario (56% [IQR, 50 to 63%]). There was a highly significant association between CPE completion and global score (R^2^ of 0.32 for *SOB* to 0.67 for *AMS*, all p < 0.001, see **SDC 12. Figure S4**).

Mean BARS scores were similar across the five axes (ranging from 5.5±1.9 for Diagnostic Processes to 6.3±1.7 for Communication. Across the four scenarios, there were no significant axes differences except for Diagnostic Process, where scores ranged from 5.1±2.0 in *Hypotension* to 5.8±1.9 in *AMS* (p = 0.006).

Overall holistic scores were 5.6±1.8 for technical and 5.8±1.8 for nontechnical performance (1-9 scale where 9 is exemplary performance) [see **SDC 10. Table S5**]. These were similar across scenarios, ranging from 5.5±2.0 in *Hypotension* to 5.9±1.8 in *AMS* (**Figure 2**). Conversely, 18.0% of *all* encounters received overall performance ratings of ≤3 (lowest bin) which varied by scenario from 14.7% for *Anxiety* to 23.5% in *Hypotension*. Overall, only 3 participants had poor bin performance scores in all 4 scenarios, <u>but 47.1% of participants scored</u> <u>≤3 on at least one scenario</u>.

**Figure 2.**
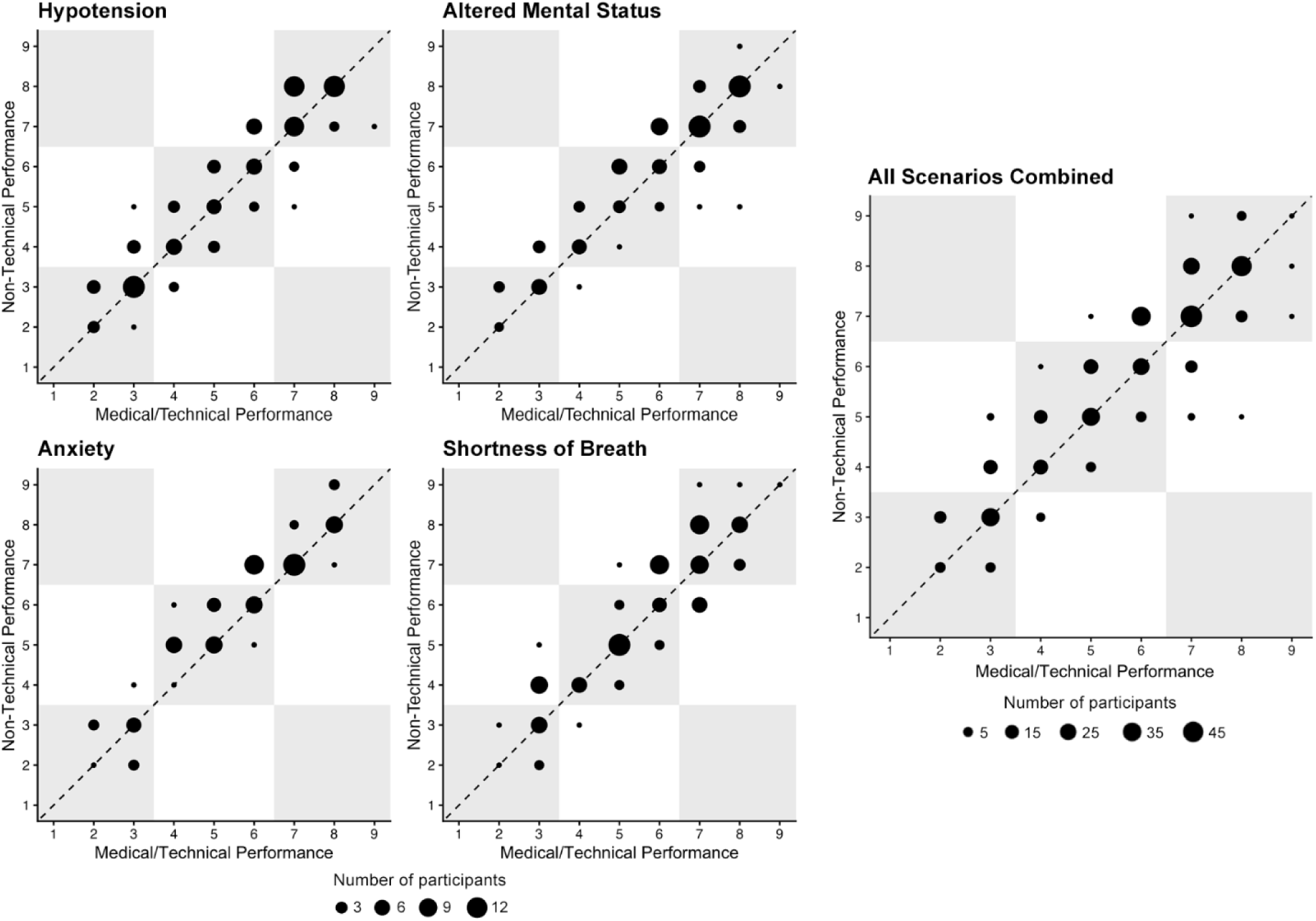
Medical/technical and behavioral/nontechnical ratings by scenario. The distribution of technical/medical performance ratings (scale 1 for worst to 9 best, see text for details) are shown against the same participant’s non-technical/behavioral performance ratings for each scenario and overall. The diameter of the dots indicates the number of participants who had this scoring pair. Most performance scores fell in the upper right third of the ‘bins’ (good to excellent performance) although up to about one-fifth of participants scored in the lowest third of the bins (poorer performance). The different scenarios show somewhat different performance score distributions (see text for details).

Seventy-eight percent of participant-scenarios were rated as having performed at Consultant Level Performance (CLP) (range 72% in *Hypotension* to 81% in *AMS* and *Anxiety)*. 40 participants achieved the CLP rating in all four scenarios; 41 achieved it in 3 scenarios. Only 2 participants did not achieve a CLP rating in any of the scenarios.

### Hypothesis 1. Effects of Training and Experience on Performance in Simulated Crises

In a binomial generalized linear mixed-effects model, the adjusted probability of completing a CPE was 0.57 (95% CI 0.54–0.61) for Juniors, 0.57 (95% CI 0.55–0.59) for BCAs, and 0.62 (95% CI 0.59–0.66) for Seniors (Two-degrees of freedom Wald test p = 0.045; **SDC-10. Table S5)**.

Aggregated across all four scenarios, on average, Seniors’ overall performance was higher than BCAs or Juniors (Seniors: 6.1±1.7, BCAs 5.6±1.8, and Juniors 5.4±1.8, p = 0.083). Similar trends were observed in both technical and non-technical performance, as well as BARS (**SDC-11. Table S5**). Average percentage of CPEs completed were highest for Seniors (62%), followed by BCAs (57%) and Juniors (56%). Conversely, 54.5% of Juniors had a rating ≤3 in at least one scenario compared to 47.5% of BCAs and 36.8% of Seniors.

In a linear mixed effects regression model for overall performance, holding all other covariates in the model constant, Seniors scores averaged 0.6 points higher than Juniors and 0.5 higher than BCAs (global p-value = 0.195). No significant differences were observed by scenario (p = 0.284) or participant sex (p = 0.647). Similar results were observed in a logistic mixed-effects regression for attainment of consultant level performance, although participants were almost half as likely to attain CLP in *Hypotension* than in *Anxiety* (OR 0.54, 95% CI 0.27 – 1.08, p = 0.083; **SDC-12. Table S6**).

The strongest evidence relevant to Hypothesis 1 was the robust <u>nonlinear</u> rather than linear association between overall performance and the continuous measure of the *combined years of training and experience* (p = 0.008) (**Figure 3**). Predicted overall performance increased from 5.5 (95% CI 4.9–6.2) at one year to a peak of approximately 6.4 (95% CI 5.9–6.8) at 5 years of combined training and experience (i.e., ∼2 years post-training). Beyond this point, performance declined gradually to a nadir of 5.0 (95% CI 4.5–5.5) at 16–17 years of total training and experience.

**Figure 3.**
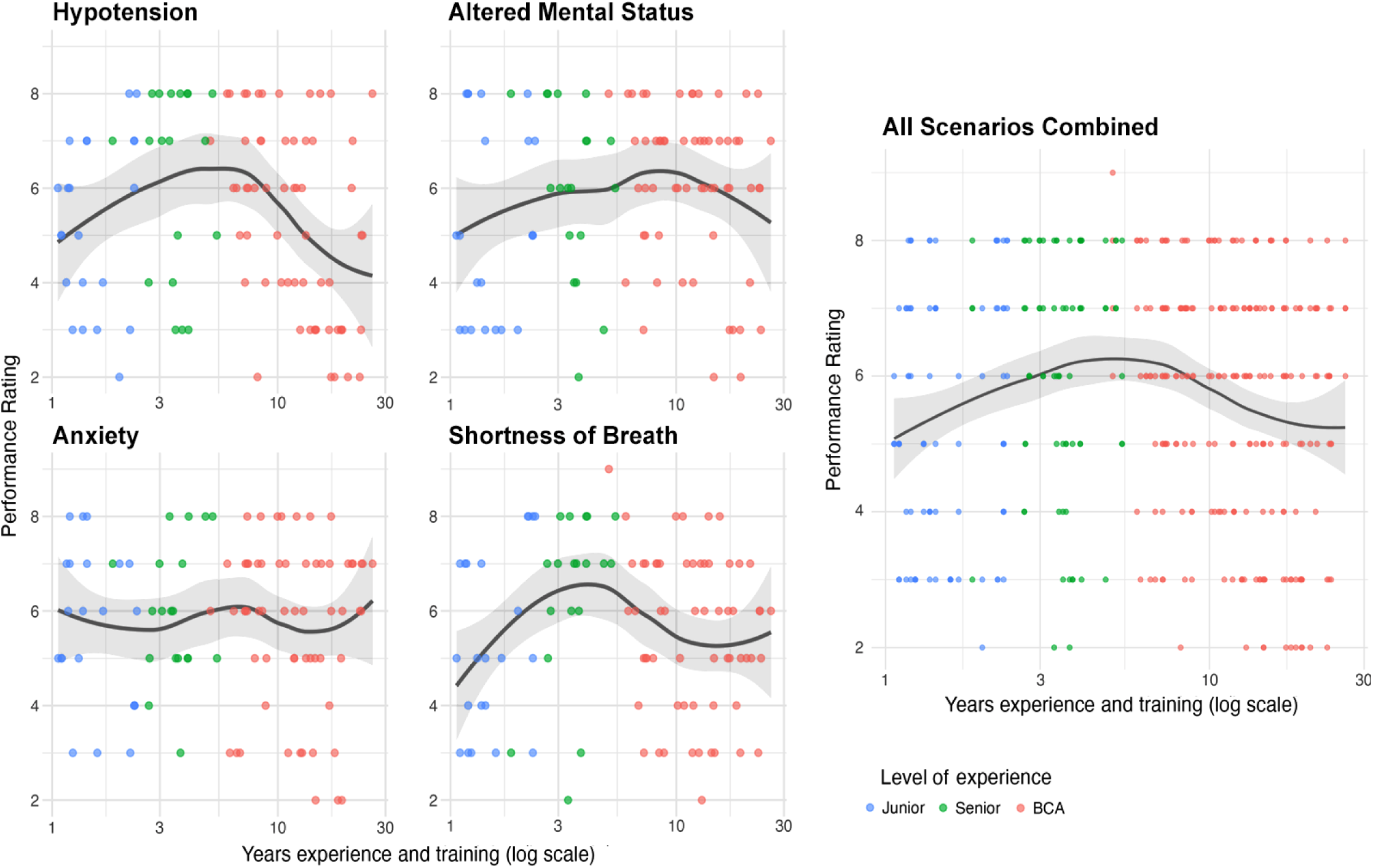
Global performance for each scenario by years of training and experience. The distribution of global ratings (same 9-point scale as above) is shown for each scenario by total years since the start of training (log scale). The dots are color coded to indicate the cohort (Junior, Senior, or BCA – see Methods for categorization criteria) of each study participant. Highest scores are generally seen for participants who have had at least 2 years of training which a fall-off in 3 of the 4 scenarios after some years in practice (especially for Hypotension/Urosepsis, the most difficult scenario).

### Hypothesis 2. Effects of Prior Crisis Management Experience on Performance in Simulated Crises

In the linear mixed effects regression model for overall performance, the global test of association was significant for crisis experience principal components (p = 0.040) but *not* for either clinical experience (p = 0.972) or simulation experience (p = 0.937) (**SDC-12. Table S6**). There was also a significant association (p = 0.015) between the crisis experience components and greater likelihood of Consultant Level Performance. Similar but non-significant trends were observed in the medical/technical performance and non-technical performance scores.

### Hypothesis 3. Identified Performance Gaps will Reiterate and Expand Prior Findings

CPE performance was strongly correlated with global scores (Spearman’s ρ = 0.72, p < 0.001) (see **SDC-13** and **SDC-14. Table S7**). Across all scenarios, a 10% increase in the proportion of CPEs marked as present was associated with a 2.3 (95% CI 2.1–2.5) point increase in global score (p < 0.001). In at least one scenario, a high proportion of participants failed to call for help (*SOB*) or engage/consult with other clinicians *(Hypotension, AMS)*, obtained advanced diagnostic tests (e.g., EKG in *SOB*, ABG or CXR in *Hypotension*, troponins in *Anxiety*), ordered POC ultrasound *(SOB, Hypotension, Anxiety*), escalated hemodynamic support (S*OB, Hypotension, Anxiety)*, or broadened their therapeutic approach (e.g., antibiotic coverage in *SOB* and *Hypotension*, stress dose steroids in *Hypotension*, treatment of serotonin syndrome in *AMS*) (**Table 3**). The failure to complete many specific CPEs was associated with lower global performance scores.

**Table 3.** CPE Completion Rates and Association with Global Performance Scores*.

| Clinical Performance Element (CPE) |  | Percent completed | General Performance Category | Association with Higher Global Performance Score |  |  |
| --- | --- | --- | --- | --- | --- | --- |
| CPE # |  |  |  | Beta | 95% CI | p-value |
| Shortness of breath (SOB) |  |  |  |  |  |  |
| 1 | Obtains a patient history AND Reviews chart/anesthesia record | 100.00% | Initial evaluation | N/A | N/A |  |
| 2 | Performs a physical exam (airway, heart-lung, OR abdominal exams) | 97.00% | Initial evaluation | -0.4 | -2.4 - 1.7 | 0.723 |
| 3 | Escalates supplemental oxygen levels | 77.20% | Initial therapy | 0.4 | -0.4 - 1.2 | 0.36 |
| 4 | Calls for clinician assistance [MD, RN, or other] | 46.50% | Seeks assistance | 0.8 | 0.1 - 1.5 | 0.024 |
| 5 | Gives intravenous fluid bolus of at least 500cc | 86.10% | Initial therapy | 1.4 | 0.4 - 2.4 | 0.005 |
| 6 | Administers a pressor (bolus) or starts an infusion | 81.20% | Initial therapy | 1 | 0.2 - 1.9 | 0.018 |
| 7 | Obtains a chest X-Ray | 51.50% | Diagnostic testing | 0.4 | -0.3 - 1.1 | 0.233 |
| 8 | Obtains an ABG | 71.30% | Diagnostic testing | 0.8 | 0.1 - 1.6 | 0.026 |
| 9 | Secures or is prepared to imminently secure the airway | 63.40% | Escalate therapy | 0.9 | 0.2 - 1.6 | 0.009 |
| 10 | Obtains a 12-lead EKG | 30.70% | Diagnostic testing | 0.4 | -0.4 - 1.1 | 0.324 |
| 11 | POC Ultrasound to evaluate cardiac, lungs, abdomen | 16.80% | Diagnostic testing | 1.1 | 0.2 - 2 | 0.014 |
| 12 | Escalates care by either calling for an ICU bed or contacting surgeon to discuss further management | 77.20% | Escalate therapy | 1.2 | 0.4 - 2 | 0.003 |
| 13 | Orders additional labs (based on reasonable diagnoses) | 61.40% | Diagnostic testing | 0.5 | -0.2 - 1.2 | 0.159 |
| 14 | Escalates hemodynamic support | 10.90% | Escalate therapy | 1.1 | 0 - 2.2 | 0.046 |
| 15 | Contact Blood Bank | 1.00% | Seeks assistance | N/A | N/A | N/A |
| 16 | Reviews and broadens antibiotic coverage | 5.00% | Escalate therapy | 0.6 | -1 – 2.2 | 0.469 |
| Hypotension |  |  |  |  |  |  |
| 1 | Obtains a patient history AND Reviews chart/anesthesia record | 99.00% | Initial evaluation | N/A | N/A | N/A |
| 2 | Performs a physical exam (airway, heart-lung, OR abdominal exams) | 86.30% | Initial diagnosis | 1.7 | 0.6 - 2.8 | 0.003 |
| 3 | Gives intravenous fluid bolus at least 500cc | 85.30% | Initial therapy | 1.5 | 0.4 - 2.5 | 0.007 |
| 4 | Administers a pressor (bolus) or starts an infusion | 98.00% | Initial therapy | N/A | N/A | N/A |
| 5 | Escalates supplemental oxygen levels | 93.10% | Escalate therapy | 1.4 | -0.1 - 2.9 | 0.067 |
| 6 | Obtains an ABG | 43.10% | Diagnostic testing | 0.6 | -0.2 - 1.4 | 0.123 |
| 7 | Obtains a 12-lead EKG | 67.60% | Diagnostic testing | -0.4 | -1.2 - 0.4 | 0.328 |
| 8 | Obtains a chest X-Ray | 48.00% | Diagnostic testing | 1.2 | 0.4 - 1.9 | **0.002** |
| 9 | Contacts urologist to discuss patient status and concerns | 38.20% | Seeks assistance | 1 | 0.3 - 1.8 | **0.008** |
| 10 | POC Ultrasound to evaluate cardiac, lungs, abdomen | 20.60% | Diagnostic testing | 1.2 | 0.3 - 2.2 | **0.01** |
| 11 | Calls for clinician assistance [MD, RN, or other] | 72.50% | Seeks assistance | 0.6 | -0.3 - 1.4 | 0.19 |
| 12 | Orders additional labs (based on reasonable diagnoses) | 55.90% | Diagnostic testing | 1.6 | 0.9 - 2.3 | **<0.001** |
| 13 | Gives additional volume | 26.50% | Escalate therapy | 1.4 | 0.6 - 2.2 | **0.001** |
| 14 | Secures or is prepared to imminently secure the airway | 87.30% | Escalate therapy | 1 | -0.2 - 2.1 | 0.101 |
| 15 | Escalates hemodynamic support | 36.30% | Escalate therapy | 1.1 | 0.3 - 1.9 | **0.005** |
| 16 | Reviews and broadens antibiotic coverage | 9.80% | Escalate therapy | 1.8 | 0.6 - 3.1 | **0.005** |
| 17 | Gives stress dose steroids | 11.80% | Escalate therapy | 1.3 | 0.1 - 2.5 | **0.033** |
| **Altered mental status (AMS)** |  |  |  |  |  |  |
| 1 | Asks anesthesiologist about patient history and possible precipitants | 99.00% | Initial evaluation | N/A | N/A | N/A |
| 2 | Orders additional labs (based on reasonable diagnoses) | 62.70% | Diagnostic testing | 0.9 | 0.2 - 1.6 | **0.011** |
| 3 | Obtains a blood glucose level | 86.30% | Diagnostic testing | 1.1 | 0.1 - 2.1 | **0.033** |
| 4 | Conducts more extensive neurological exam | 61.80% | Initial diagnosis | 1.8 | 1.2 - 2.4 | **<0.001** |
| 5 | Attempts to dissuade/prevent confederate anesthesiologist from giving naloxone and/or flumazenil | 49.00% | Other | 0.6 | -0.1 - 1.3 | 0.109 |
| 6 | Articulates need to broaden the differential diagnosis (beyond the common diagnoses in this situation) | 66.70% | Broaden DDx | 2 | 1.3 - 2.6 | **<0.001** |
| 7 | Administers sedative/hypnotic | 70.60% | Escalate therapy | 1.8 | 1.1 - 2.5 | **<0.001** |
| 8 | Considers either serotonin syndrome OR neuroleptic malignant syndrome in differential diagnosis. | 60.80% | Broaden DDx | 1.4 | 0.7 - 2.1 | **<0.001** |
| 9 | Orders administration of cyproheptadine (Periactin™) or chlorpromazine (Thorazine™) | 10.80% | Escalate therapy | 2.2 | 1.1 - 3.4 | **<0.001** |
| **Anxiety** |  |  |  |  |  |  |
| 1 | Obtains a patient history AND Reviews chart/anesthesia record | 99.00% | Initial evaluation | N/A | N/A | N/A |
| 2 | Performs a physical exam (airway and heart-lung exam) | 36.3% | Initial evaluation | 0.5 | -0.2 -1.1 | 0.168 |
| 3 | Obtains a 12-lead EKG | 87.30% | Diagnostic testing | 2.5 | 1.6 - 3.3 | **<0.001** |
| 4 | Recognizes and articulates myocardial ischemia | 75.50% | Broaden DDx | 2.6 | 2 - 3.1 | **<0.001** |
| 5 | Orders additional labs (based on reasonable diagnoses) | 36.30% | Diagnostic testing | 0.6 | -0.1 - 1.3 | 0.079 |
| 6 | Administers supplemental oxygen | 96.10% | Initial therapy | 1.8 | 0.2 - 3.5 | **0.029** |
| 7 | Escalates supplemental oxygen levels | 59.8% | Escalate therapy | 1 | 0.3 - 1.6 | **0.003** |
| 8 | Orders cardiac enzymes | 47.10% | Diagnostic testing | 1.5 | 0.9 - 2.1 | **<0.001** |
| 9 | Administers a pressor (bolus) | 43.10% | Escalate therapy | 0.8 | 0.2 - 1.5 | **0.011** |
| 10 | Calls cardiology | 72.50% | Seeks assistance | 2.6 | 2.1 - 3.1 | **<0.001** |
| 11 | Administers additional pressors (bolus or infusion) | 15.70% | Escalate therapy | 1.3 | 0.5 - 2.2 | **0.003** |
| 12 | Asks for POC ultrasound to evaluate cardiac function | 14.70% | Diagnostic testing | 0.6 | -0.3 - 1.5 | 0.195 |

\* We analyzed which CPEs were associated with overall performance ratings using a linear regression model for each CPE. The estimated effect (Beta) for each model relates the average difference in overall performance between those who completed the CPE compared to those who did not. Empty cells for regression results denote that no model was fit due to extremely high or low completion rates.

## Discussion

In this multi-site simulation-based study, anesthesiologists with more real-world or simulated experiences in crisis situations outperformed their peers, reinforcing the importance of consistent experiential learning in developing and maintaining crisis response skills. Notably, rated performance peaked at 5 years of training plus experience (i.e., about the time of initial board certification) and nearly half of participants demonstrated ineffective crisis management in at least one scenario, consistent with the MOCA study^9^ and prior simulation-based studies.^10,26,27^ These findings underscore the value of structured ongoing training, particularly for anesthesiologists who do not have frequent real-world exposure to clinically challenging situations.

This multi-scenario structure allowed for examination of performance across event types, acknowledging that proficiency in one crisis does not guarantee competence in others, which was reflected in the scoring differences between scenarios. Further validity evidence is provided by the lower scores of Juniors than Seniors or BCAs. The reproducibility of previous findings, particularly poorer simulation performance associated with greater time elapsed since the end of training,^9^ reinforces the validity of using simulations to evaluate real-world crisis management capabilities.^9,28,29^ While this study focused on anesthesiologists, scenarios were designed to evaluate competencies needed to manage acute clinical events. Thus, the methods and findings could be relevant to other acute care clinicians. The study population’s holistic performance was generally scored higher than in our prior simulation-based assessment study,^9^ a finding possibly attributable to scenario difficulty, participant ability, or rater stringency.

This study offers several insights into the determinants of performance. First, anesthesiologists who rarely encountered high-acuity situations in their routine practice were more likely to perform poorly, suggesting that limited cognitive rehearsal and skill decay since the completion of residency training contribute meaningfully to crisis management deficits. Second, despite the intuitive expectation that clinical practice environment/training might play a role, we found no performance differences between academic and community anesthesiologists or by subspecialty, again suggesting that individual training and experience is a primary driver of crisis management readiness. Finally, the frequent failure to perform expected clinical behaviors such as calling for help, timely diagnosis broadening, and early hemodynamic escalation identify where targeted cognitive and behavioral training may be especially beneficial. Despite differing clinical scenarios, many performance gaps identified in this study replicate our previous findings.^9,20^ Both studies demonstrated communication deficiencies (e.g., to communicate effectively with colleagues), failure to refine/expand diagnosis (e.g., not ordering additional tests, fixation upon an unlikely diagnosis), failure to escalate treatment sufficiently, and technical/knowledge deficiencies (e.g., failure to broaden antibiotic coverage in sepsis, use of steroids for steroid dependent patients, recognition and treatment of the serotonin syndrome, and hypovolemia as cause of shock in TRALI).

### Limitations

This study has limitations. The sample size was modest, representing only four geographic regions. Voluntary participation may have introduced selection bias. Our study population included more females than the national anesthesiologist average, although there were no sex-based performance differences. Participants received tangible incentives, which could have biased sampling and performance.

Although we collected extensive data on training and practice attributes, unmeasured factors, practice heterogeneity, or dimensionality reduction may have influenced our findings. Anesthesiologists more than ∼25 years out of training were excluded, limiting our ability to evaluate late career clinicians. COVID-related modifications could have affected both data collection and analysis. Scenario delivery may have varied across sites, although we went to appreciable effort to reduce this, and study site was not a significant covariate in the regression analyses. Performance in simulation may differ from real clinical behavior, though prior evidence supports simulation’s predictive validity. Simulations cannot perfectly emulate participant’s actual practice setting (e.g., the people and technological support structures that evolve to optimize team performance) so that individual evaluations may not reflect actual real-world behavior. The absence of performance differences after adjusting for prior simulation experience is consistent with earlier studies,^30,31^ and may reflect the ubiquitous use of simulation-based training in residency in recent decades.

Prior work suggested that at least eight diverse scenarios would be required to accurately assess individual *competence*.^32^ In the present study, each participant did four scenarios. Two, the *SOB* and *Hypotension* scenarios, were similar in expected management of acute cardiorespiratory decompensation – and performance was similar between them. The other two scenarios were less difficult and presented nontechnical challenges. Participants had to do the study cases ‘solo’ whereas only 23% of our BCAs were solo practitioners. However, the cases were all pre- and post-operative scenarios, common situations that anesthesiologists manage without help from colleagues.

### Implications

These findings have important implications for ongoing competency assessment, particularly for anesthesiologists who experience prolonged periods without managing critical events. The well-documented limitations of self-assessment, general laxity of peer oversight, and the absence of mandated skills assessments after primary certification make it unlikely that clinicians will independently identify or remediate their own gaps in skills and knowledge without specific mechanisms to help them do so. While written and oral examinations assess knowledge and judgment, they may not reliably capture the dynamic cognitive, behavioral, and team-based skills required during real crises. Simulation-based assessment has been incorporated into the ABA’s primary certification processes,^19^ including team-based skills, and offers a unique opportunity to evaluate these skills directly, to identify performance patterns that may benefit from structured remediation, and to advance the skills of all anesthesiologists going forward.

In this study, we identified several categories of gaps (see **Table 3** and **SDC-13. Table S7**), including issues with ordering additional tests, escalating therapy, seeking assistance from colleagues, and broadening the differential diagnosis. Evidence of gaps was seen in the prior MOCA study across all these categories except differential broadening.^9,20^ A key component of anesthesiologists’ lifelong learning and maintenance of clinical competencies is their ability to self-assess learning needs and engage in self-targeted educational activities. However, self-assessment often fails to identify competency gaps, hindering effective self-improvement.^33,34^ This is compounded by the lack of mandatory competency assessments after board certification. Traditional written exams, which focus largely on medical knowledge (i.e., knows what to do), are poor predictors of real-world performance (i.e., does it appropriately). The consistent identification of important performance gaps around diagnosis, management, and teamwork during acute care events, which persist or even worsen in experienced practitioners, point to the need for targeted efforts to improve skills through adult learning modalities such as the ASA-sponsored MOCA simulation courses.^35^

### Conclusions

This rigorous multi-site study highlights substantial variability in crisis management performance that appears context (event type) dependent. Clinicians who are more than 10 years out from training show degraded ability to manage challenging events. Practices or ongoing crisis management training that provides opportunities to manage challenging cases and events appears to mitigate performance degradation. Our findings also demonstrate the value of high-fidelity simulation as an assessment tool, revealing cognitive and behavioral patterns that help identify anesthesiologists who would benefit from targeted remediation or additional support.

## Supporting information

SDC 1-14

## Data Availability

All data produced in the present study are available upon reasonable request to the authors

## Contributors

1. John (Jack) Boulet, PhD was an active Co-Investigator and contributor early in this project during study design and initial implementation but did not meet all of the International Criteria for authorship.
2. Ann Harman, PhD, Chief Assessment Officer of the American Board of Anesthesiologists, was an active Co-Investigator and contributor to this project during project development and study conduct. She chose not to meet all of the International Criteria for authorship.
3. Steve Howard, MD, Professor (retired) of Anesthesiology, Perioperative and Pain Medicine Stanford University (Palo Alto, CA), was an active Co-Investigator and contributor to this project during project development and study conduct but retired in mid-2025 and chose not to meet all of the International Criteria for authorship.
4. Laura Militello, CEO of Applied Decision Science (Dayton, OH), was an active Co-Investigator and contributor to this project during project development and study conduct. She did not contribute the analysis, interpretation, or writing of this manuscript and thus failed to meet all of the International Criteria for authorship.
5. Randolph H. Steadman, MD, Professor and Chair of Anesthesiology (retired) at Weill Cornell Medical College, was an active Co-Investigator and contributor to this project during project development and study conduct but chose not to meet all of the International Criteria for authorship.

## Acknowledgements

We acknowledge Russ Beebe for his exemplary graphic design skills on numerous products throughout this project. We also acknowledge Amy Zipp and Mark Hughes for their administrative and logistical support.

IMPACTS Subject Matter Expert (SME) Panel:

Lillian Emlet, MD, MS (Emergency Med/ICU, UPMC, Pittsburgh, PA), Rosemarie Fernandez, MD (Emergency Med, Univ. Florida), Robert Gaiser, MD (Anesthesiology, Yale Univ.), Michele Gonzalez, PhD (CRNA, Univ. Arkansas), Ken Johnson, MD, MS (Anesthesiology, Univ. Utah), Joan Kinniry, MSN, (CRNA, Univ. Pennsylvania), Ed Lee, MD (Hospital Medicine, UCLA, CA), Virginia (Chris) Muckler, DNP (CRNA, Duke Univ., NC), David Murray, MD (Anesthesiology, Washington Univ., St. Louis, MO), Alexis Nicpon, MSN (PACU nurse, NW Central DuPage Hospital, IL), Manny Pardo, MD (Anesthesiology, UC San Francisco, CA)

## Video Raters

Lee-Lynn Chen, MD (UCSF), Brian Kaufman, MD (New York Univ.), Vikas Kumar, MD (Augusta, GA), Rob Morgan, MD (MUSC), Chris Nichols, MD (Univ. Colorado Children’s Hospital), Margaret O’Donoghue, MD (SUNY Albany), Louise Wen, MD (Dartmouth).

## Study site personnel

Doerthe Adriana Andreae, MD, PhD (Univ. Utah; Formerly at Penn State), Malena Agyemang, PhD (Penn State), Louise Answine, MS (PSU), Cheryl Delsega, PhD, NP (PSU), Denise Derosa, RN, MSN (Univ. New Mexico), Jonathan Eldridge, PhD (Univ. New Mexico), Sean Leadem, PhD (Univ. Pittsburgh), Amanda Lowe, PhD (Univ. Pittsburgh), Ann Miller (VUMC)

## Prior Presentations

Portions of the IMPACTS study have been presented in part at the following scholarly meetings: Society for Education in Anesthesiology (Philadelphia, PA, April 21, 2024), Euroanaesthesia (Munich, FRG, May 26, 2004), Society for Simulation in Europe (SESAM, Valencia, ES, June 24, 2025), HFES International Symposium on Human Factors and Ergonomics in Healthcare (Toronto, Canada, March 31, 2025), Healthcare Ergonomics and Patient Safety (HEPS, Dublin, Ireland, June 19, 2025), and the International Anesthesia Research Society (Montreal, Canada, May 1, 2026).

## Competing Interests

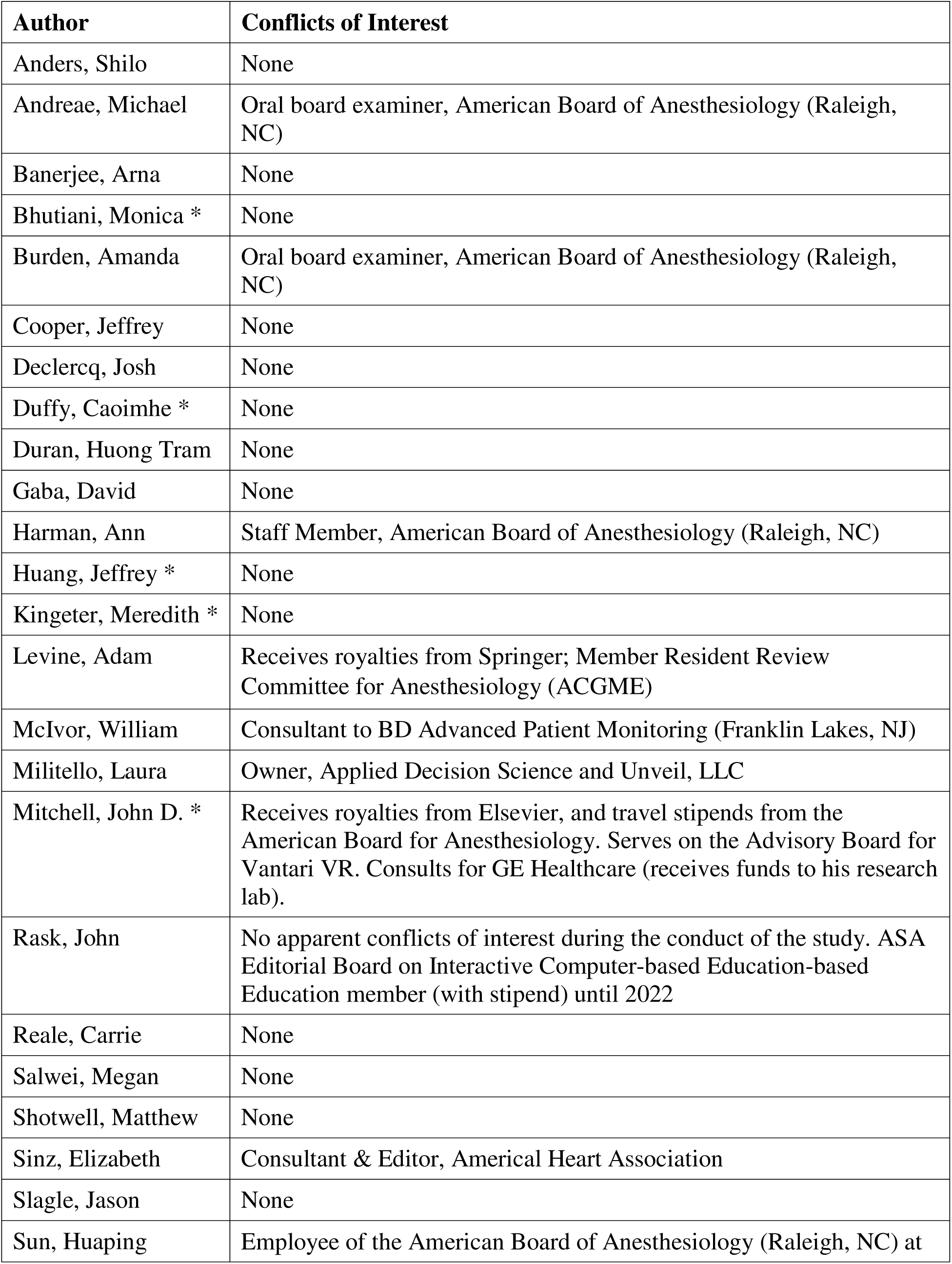
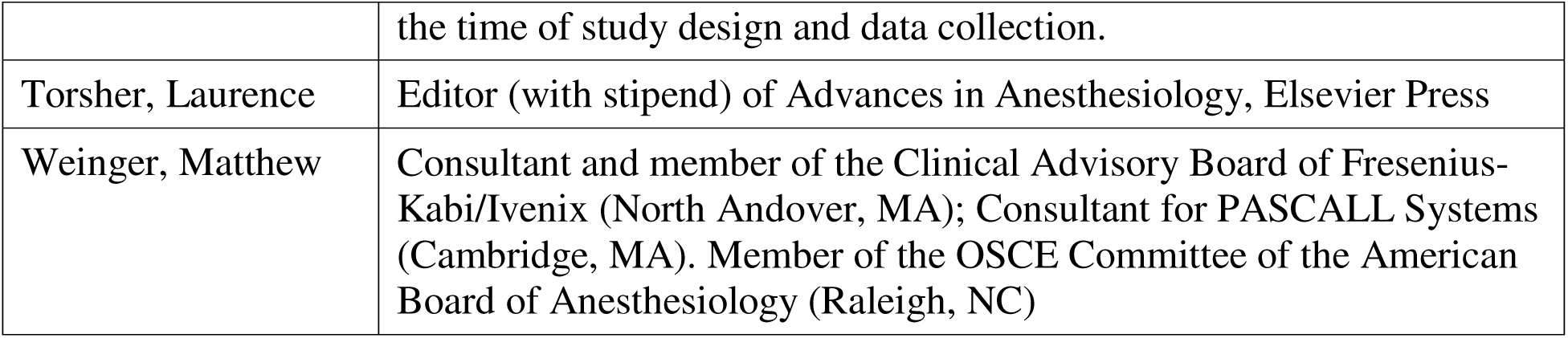

## Abbreviations

ABA: American Board of Anesthesiology
ASA: American Society of Anesthesiologists
BARS: Behaviorally Anchored Ratings Scale
BCA: Board-Certified Anesthesiologist
CI: Cognitive Interview
CME: Continuing Medical Education
CPE: Clinical Performance Element
CRM: Crisis Resource Management
IRR: Inter-Rater Reliability
Junior: An anesthesiology resident who has had at least six months of clinical anesthesiology specific training but no more than 18 months (or has sat for the CA-2 ABA In-training-exam.
MOCA™: Maintenance of Certification in Anesthesiology (by the ABA)
MOCA-Sim: The MOCA Simulation Study conducted previously by the authors
PCA: Principal Component Analysis
Senior: An anesthesiology resident or fellow who has had at least 2.5 years of clinical anesthesiology specific training but has not yet been ABA certified.
SEP: Standardized Embedded Participant (e.g., standardized clinicians or patients)
SME: Subject Matter Expert

