## Supplementary material for "Proximity to primary board certification and prior crisis management experience are associated with anesthesiologists’ performance in high-fidelity acute care scenarios": SDC 1-14

#### SDC-1. Figure S1. CONSORT diagram

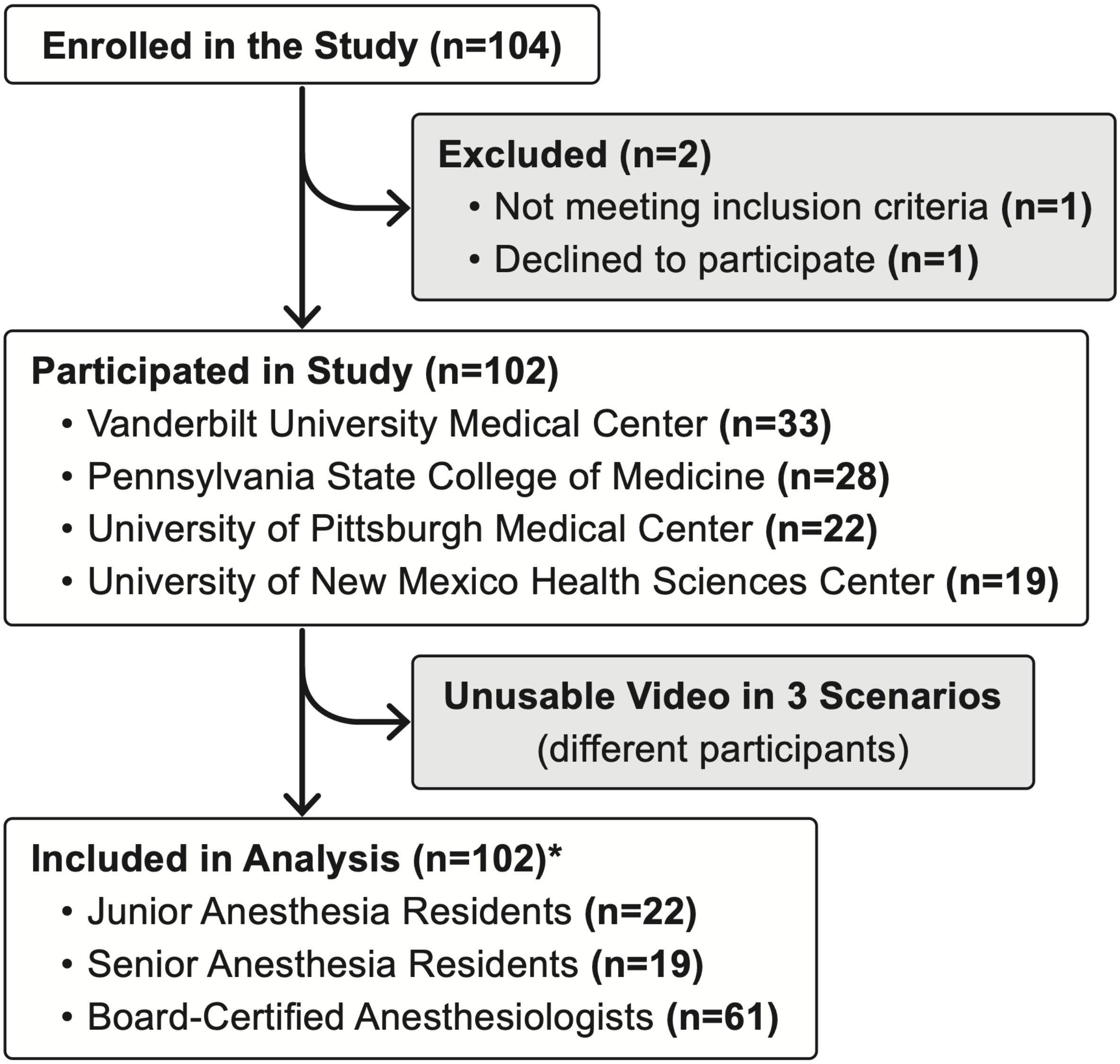


* Three individual participants had 1 unusable video.

#### SDC 2. Demographic eSurvey Questions*

* Every participant completed all 7 modules. However, the survey used branching logic so no participant answered every question and most participants answered different questions. Some of the branching logic is shown by the indented questions which were only asked if the participant answered YES to the preceding question. In the analysis, some answer options were combined for simplicity.

##### Module 1 – General Demographics and Clinical Training

1. Enter your current age. Numeric (years)
2. Enter your biological sex (M / F / Other / Decline to answer)
3. Choose the region in which you do most of your clinical work …
   - Pacific - CA, OR, WA, AK, HI
   - West - AZ, CO, ID, MT, NM, NV, UT, WY
   - Midwest - KS, IA, MN, MO, ND, NE, SD
   - West South Central - AR, LA, OK, TX
   - East North Central - IL, IN, MI, OH, WI
   - East South Central - AL, KY, MS, TN
   - South Atlantic - DE, FL, GA, MD, NC, SC, VA, WV
   - Middle Atlantic - NY, NJ, PA
   - New England - CT, MA, ME, NH, RI, VT
4. Do you use hearing aids when doing clinical work? (Y/N)
5. Do you wear corrective lens (glasses or contacts) when doing clinical work? (Y/N)
6. Are you of Hispanic/Latin ethnicity? (Y/N)
7. Are you a resident in training? (Y/N)
8. Where are you in your residency (CA1, CA2, CA3)
9. Level of training (CA1, CA2, CA3, Post-residency)
10. Are you in training to complete a subspecialty fellowship? (Y/N)
11. Are you Board Certified in Anesthesiology? (Y/N)
12. Are you currently enrolled in MOCA? (Yes, No, I currently have a lifetime ABA certification and do not need to re-certify, No, I'm not that far along in my career yet)
13. Are you a foreign medical graduate? (Y/N)
14. Name of anesthesiology training program? (free text)
    a. Start year?
    b. Expected graduation date?
15. Total years of clinical anesthesiology training in the United states? (Numeric)
16. Did you complete any non-anesthesia residencies (Y/N)
17. Are you currently in anesthesiology-related fellowship training? (Y/N)

a. What type of Fellowship Training are you currently in? (free text)
b. Expected date of completion

18. Please tell about your fellowship training in the United States (free text)
19. Have you completed anesthesiology specific fellowship training? (Y/N)
    a. What type of fellowship training did you complete? (free text)
    b. Date of completion?
20. Did you complete any other Fellowship training in any specialty? (Y/N)
    a. What other Fellowship training in any specialty did you complete?
21. Have you ever practiced another healthcare-related profession instead of medicine?
22. Are you/have you ever been active duty military? (Y/N)
23. Do you have any other advanced degree or certification? (Y/N)
24. Do you hold active certification(s) from any other board certifications? (Y/N)
    a. Which one(s)? (free text)
25. Are there any questions, in this module, that you did not understand or answers you wish to clarify? (Y/N)
    a. What questions do you need clarified or answers you wish to clarify? (free text)

#### Module 2 – Clinical Experience and Practice Attributes

1. Are you currently a resident or fellow? (Y/N)
2. How many years after completing your residency training have you actively practiced as an anesthesiologist (Subtract any time when you were not actively practicing)? (numeric)
3. Identify any leadership roles (indicate all that apply) you serve in your current institution(s) or organization(s). (narrative text)
4. Please indicate all of the following in which you consider that you currently have specialty/subspecialty expertise (this would include either special training or experience beyond that of the average anesthesiologist)
   – Regional Anesthesia/Acute Pain
   – Ambulatory/Outpatient
   – Pediatric Anesthesia
   – Teaching (beyond ordinary case supervision or occasional talks)
   – Cardiothoracic Anesthesia
   – General Operative Anesthesia
   – Pain Medicine
   – Administration
   – Critical Care Medicine
   – Thoracic Anesthesia

- Trauma Anesthesia
- Neuro Anesthesia
- Obstetric Anesthesia
- Vascular Anesthesia
- Cardiac Anesthesia
- Research
- Hospice and Palliative Care
- Sleep Medicine

5. What is the average total number of hours you work each week? (numeric)
6. What is your average work day length (hours)? (numeric)
7. How many days per week (on average) do you work more than 12 hours continuously? (numeric)
8. How many days per week (on average) do you work more than 18 hours continuously? (numeric)
9. How many hours each week do you spend doing clinical work (including hands on anesthesia care, direct supervision of nurse anesthetists, residents, or anesthesia assistants)? (numeric)
10. Thinking about your average work week, how do you spend your time on the following activities? The total should add up to 100%.
    - Hands-on clinical work (doing your own cases)
    - Hands-on clinical work (supervising other clinicians)
    - Administrative activities
    - Education (excluding direct clinical supervision)
    - Research and other scholarly activities
    - Other:
11. Do you have night or weekend call responsibilities? (Y/N)
    a. On average, how many nights per month are you on the following:
       - In-House Night Call
       - Pager night call in which you sometimes need to come back to the hospital
       - Pager night call in which you handle issues by phone and almost never need to come back to the hospital
12. In the average week, how many hours after 5 pm do you work clinically?
13. On average, how many weekend days per month do you do in-house call? Include days when you are on beeper call but must come back to the hospital to do a case. Do not include pager call that does not require you to come back to the hospital.
14. Are there any questions, in this module, that you did not understand or answers you wish to clarify? (Y/N)
    a. What questions need clarification or answers you need to clarify? (free text)

#### Module 3 – Clinical Practice Attributes

**Section Header:** In this section, we will learn about the nature of your clinical practice, the kind of clinical sites at which you work, and the resources available to you at those sites. Many anesthesiologists work at more than one clinical site, and if that is the case for you, we want to learn about the practice and resources available at the two sites that encompass the majority of your practice.

We define a clinical site as a place that may have many anesthetizing locations, like a main operating room, ambulatory surgery suite, or gastrointestinal lab, with unique types of cases, surgeons, and patient populations, but all those locations have basically similar clinical processes and policies, staff, equipment, supplies, and the like. The clinical site has a culture and an anesthesiologist could move comfortably among the various anesthetizing locations and feel at home.

In contrast is a stand-alone surgical center that is located several miles away from the main hospital. Even though it may be part of the same healthcare system, the surgi-center is more likely to have different clinical processes and policies, staff, equipment, supplies, and patient populations (e.g., healthier patients having different kinds of surgeries) than the main hospital. We consider this example surgi-center as a different clinical site."

1. How many clinical sites do you work at?
2. Site 1 name (narrative text)
3. Site 2 name (narrative text)
4. For each site, choose the best descriptor:
   a. Tertiary Care, Academic Medical Center
   b. Tertiary Care, Non-Academic Medical Center
   c. Community Medical Center (may have an Academic Affiliation)
   d. Regional Medical Center
   e. Standalone Ambulatory Center
   f. Surgeon's office
   g. Other
5. Which do you consider your 'Primary practice site' (i.e., where you do most of your clinical work)?
6. How many miles is [site1] from [site_2]?

*Site 1 and Site 2 are both asked where see [work-site] below…*

7. How many years have your worked at [work_site]?
8. How many days per week on average do you work at [work_site]?
9. On average, how long, in hours, is your typical workday at [work_site]?
10. What percentage of your clinical work is done at [work_site]?
11. During clinical work at [work_site], indicate the frequency with which you:
    a. Do your own cases?

   b. Medically direct experienced anesthesia providers (CRNA or AA)
   c. Supervise providers in-training (residents, SRNAs, SAAs)
   d. Supervise (>1:5) experienced anesthesia providers (CRNA)
12. How many codes have you managed at [work_site] in the last 3 months?
13. How frequently are you actively involved in hands-on crisis management drills like mock codes at [work_site]?
14. Do you have access to emergency manuals or checklists that provide detailed guidance on the management of clinical emergencies at [work_site]?
   a. How frequently do you use emergency manuals or checklists in patient care? (never, sometimes, frequently)
15. Which of the following resources do you have access to at [work_site]: (Check all that apply)
   a. Anesthesia technician
   b. Code cart
   c. MH box or cart
   d. Difficult airway box or cart
   e. TEE machine and probe
   f. Blood bank
   g. Cardiac catheterization laboratory
   h. Interventional radiology
   i. Cardiopulmonary bypass machine
   j. ECMO machine
   k. Intensive Care Unit
   l. Emergency Room
   m. Hemodialysis
16. Which of the following personnel are immediately available in an anesthesia emergency at [work_site] during regular work hours: (Check all that apply)
   a. Another experienced anesthesiologist
   b. An inexperienced anesthesiologist (i.e., resident)
   c. An experienced non-MD anesthesia professional (i.e., CRNA or AA)
   d. An inexperienced non-MD anesthesia professional (i.e., SRNA or SAA)
   e. Code or rapid response team
   f. Intensive care physician
   g. Intensive care nurse practitioner
   h. Emergency room physician
   i. Trauma or general surgeon
   j. None of the above
17. How long would it take (in minutes) for the first individual indicated above to respond? If you're not sure please state, 'Don't know'.

18. Which of the following personnel are immediately available in an anesthesia emergency at [work_site] on weekends: (Check all that apply)
    a. Another experienced anesthesiologist
    b. An inexperienced anesthesiologist (i.e., resident)
    c. An experienced non-MD anesthesia professional (i.e., CRNA or AA)
    d. An inexperienced non-MD anesthesia professional (i.e., SRNA or SAA)
    e. Code or rapid response team
    f. Intensive care physician
    g. Intensive care nurse practitioner
    h. Emergency room physician
    i. Trauma or general surgeon
    j. None of the above
19. How long would it take (in minutes) for the first individual indicated above to respond? If you're not sure please state, 'Don't know'.
20. At [work_site], how of your many patients for whom you provide direct anesthesia care are: (none, some, many)
    a. ASA 3 or higher
    b. Acutely unstable (e.g., come directly to the OR from the ED or the ICU in unstable condition as a 4E or 5E)
    c. Emergencies like acute appendicitis or open fractures who are hemodynamically stable (2E or 3E)
21. Identify any factors at this site that make work there especially clinically challenging (e.g., high turnover of OR staff, old equipment, etc.). If you can't think of any, mark it as 'none'.
22. Are there any other characteristics at [work_site] that make the occurrence of unexpected acute events more common than in typical practice of elective surgery? (Y/N)
    a. Please describe these characteristics (narrative text)
23. Please indicate how often on average you spend time on all of the following activities at [work_site): (never, sometimes, frequently)
    a. Administration
    b. Ambulatory/Outpatient
    c. General (e.g. general, orthopedics, ENT, plastics, urology) non-complex (ASA 1-3) ADULT cases
    d. General (e.g. general, orthopedics, ENT, plastics, urology) non-complex (ASA 1-2) PEDIATRIC cases
    e. Pediatric cases for sick patients (ASA 3+)
    f. Patients with a BMI >40
    g. Offsite cases (GI, Neuro IR, others)
    h. Cardiac (open or endovascular, including EP)

   i. Critical care
   j. Hepatic (major, non-transplant)
   k. Neuro anesthesia (intracranial)
   l. Spine (major, >3 levels or major instrumentation)
   m. Thoracic (one-lung ventilation)
   n. Transplant (hepatic, renal, pancreas)
   o. Trauma, acute
   p. Vascular (major)
   q. Obstetrics, lower risk (ASA 1-2)
   r. Obstetrics, high risk (ASA 3-4)
   s. Regional anesthesia/acute pain
   t. Pain medicine – chronic
   u. Research
   v. Other

24. In a typical week, how often do you perform these procedures personally? (never, sometimes, frequently)
   a. Arterial line
   b. Central line
   c. Pulmonary artery catheter
   d. Perioperative TEE or TTE
   e. Point of care ultrasound
   f. Non-invasive cardiac output monitoring
   g. Cerebral monitoring
   h. Electrophysiological monitoring
   i. Awake fiberoptic intubation
25. In a typical week, how often do you supervise an **inexperienced** provider (or trainee) do these procedures? (never, sometimes, frequently)
   a. Arterial line
   b. Central line
   c. Pulmonary artery catheter
   d. Perioperative TEE or TTE
   e. Non-invasive cardiac output monitoring
   f. Any form of cerebral monitoring
   g. Awake fiberoptic intubation
26. In a typical week, how often do you supervise an **experienced** provider (or trainee) do these procedures? (never, sometimes, frequently)
   a. Arterial line
   b. Central line

   c. Pulmonary artery catheter
   d. Perioperative TEE or TTE
   e. Non-invasive cardiac output monitoring
   f. Any form of cerebral monitoring
   g. Awake fiberoptic intubation
27. Which of these sites (1 or 2) do you think is the most challenging clinically?
28. In the last 10 years, have you worked at another site or type of practice that was significantly more challenging?
29. Did you practice at this site for more than 6 months?
30. Do you frequently work at other work sites other than [work_site]?
31. Are there any questions, in this module, that you did not understand or answers you wish to clarify? (Y/N)
   a. What questions do you need clarified or answers you wish to clarify? (free text)

#### Module 4 – Clinical Practice Attributes

**Section Header:** In this section, we want to gauge the frequency of 'challenging situations' you face in your practice.

We define a 'challenging situation' as having at least one, but typically more, of the following attributes:

- Unexpected significant clinical event or situation (like an MI, abrupted placenta, or anaphylactic reaction)
- Significant expected event or situation that was not responsive to typical interventions (like refractory hypotension from an angiotensin receptor blocker)
- Sick patient (having to anesthetize a patient with a threatened limb who is in heart failure, for example)
- Complex surgical procedure or significant unexpected surgical event (e.g. bleeding)
- Use of complex technology or technique (e.g. CPB, ECMO, one-lung ventilation)

AND

- Taking at least 15 minutes to transpire

1. How many such "challenging situations" have you either managed yourself, supervised someone else managing, or assisted a colleague in managing in the LAST MONTH?
2. How many such "challenging situations" have you either managed yourself, supervised someone else managing, or assisted a colleague in managing in the LAST YEAR?

*If 0 in the above question:*

3. How many years has it been since you managed one yourself, supervised someone else managing one, or assisted a colleague in managing one?
4. Are there any questions, in this module, that you did not understand or answers you wish to clarify? (Y/N)
   a. What questions do you need clarified or answers you wish to clarify? (free text)

#### Module 5 – Crisis Management Training and Simulation Experience

5. Are you currently certified in:
   a. ACLS
   b. PALS
   c. ATLS
6. Have you completed a MOCA Simulation Course at an ASA Endorsed Center?
7. Have you taken a hands-on anesthesia or critical care simulation course (e.g., Anesthesia-centric-Advanced Cardiac Life Support (A-ACLS)) at an academic meeting or simulation center?
8. Have you taken any other type of simulation-based course?
   a. Did the course include teamwork and crisis management?
9. Have you had any other formal training in clinical crisis management?
10. Have you had any other formal training in team based clinical skills that involved active role playing?
11. Have you worked with the ASA's SimStat™ screen-based simulation training modules?
12. Are you a trainer in any of the above activities?
13. Other relevant training to crisis management and teamwork?
14. Are there any questions, in this module, that you did not understand or answers you wish to clarify? (Y/N)
    a. What questions do you need clarified or answers you wish to clarify? (free text)

#### Module 6. Hazardous Attitudes Survey

1. My success as an anesthesiologist is entirely attributable to my hard work and ability. (self-confidence)
2. When doing anesthesia for trauma, I often worry about getting into trouble. (worry/anxiety)
3. If my anesthetic management depends on a specific piece of equipment functioning, I worry about having to complete the anesthetic safely if that equipment fails. (worry/anxiety)

4. I can learn any clinical skill, if I put my mind to it. (self-confidence)
5. I like to do challenging procedures. (macho)
6. I really hate delaying cases. (impulsive)
7. I really like to do challenging cases (macho)
8. I really worry about having to abort a case in the middle. (worry/anxiety)
9. If the patient is medically unstable, I will not delay the case unless there is a major problem. *reverse scored* (impulsive)
10. I always worry about technical errors (like picking the wrong drug or making the wrong diagnosis) and complications when conducting an anesthetic. (worry/anxiety)
11. I am basically an impatient anesthesiologist. (impulsive)
12. I like high-risk cases. (macho)
13. The OR administration is more interested in controlling physician behavior in the OR than providing quality service. (anti-authority)
14. Sometimes I feel I have very little control over what happens to the patient. (resignation/ external loc)
15. In a tight situation, I trust fate. (resignation/ external loc)
16. I really worry about needlestick injuries. (worry/anxiety)
17. Sometimes I feel like the patient's outcome is set before I even start. (resignation/ external loc)
18. The thoroughness of my preoperative plan mostly determines the likelihood of my having a problem during the case. (self-confidence)
19. The OR scheduling desk is more of a hindrance than a help. (anti-authority)
20. When I am in a tough spot, I figure if I make it, I make it and if I do not, I do not. (resignation/external loc)
21. I like to do difficult procedures. (macho)
22. If you want to protest a license suspension by the state medical licensing authority, the odds are stacked against you. (impulsive)
23. In anesthesiology- what will be will be. (resignation/external loc)
24. In general, I find the OR scheduling desk to be very helpful. *reverse scored* (anti-authority)
25. If I have done something wrong in the OR, I will report it because someone will report me anyway. (self-confidence)
26. A successful anesthetic is solely the result of good planning and good execution. (self-confidence)
27. Most OR administration rules do not promote safety. (anti-authority)
28. I like to do unusual cases. (macho)
29. Are there any questions, in this module, that you did not understand or answers you wish to clarify? (Y/N)
    a. What questions do you need clarified or answers you wish to clarify? (free text)

#### Module 7. Clinician Burnout (Two-item version)

1. I feel burned out from my work. (Never, A few times a year or less, Once a month or less, A few times a month, Once a week, A few times a week, Every day)
2. I've become more callous toward people since I took this job. (Never, A few times a year or less, Once a month or less, A few times a month, Once a week, A few times a week, Every day)

#### SDC 3. Table S1. Panel of Subject Matter Experts (SMEs) used in the Scenario Design and CPE Development Process

| SME's Name § | Degrees | Discipline | Current Titles | Current Primary Affiliation |
|---|---|---|---|---|
| Lillian Emlet | MD, MS, FCCM | Critical Care Medicine, Emergency Medicine | Associate Professor of Critical Care Medicine; Associate Program Director, Internal Medicine Critical Care Fellowship. | University of Pittsburgh Medical Center (UPMC) and VA Pittsburgh Healthcare System, Pittsburgh, PA |
| Rosemarie Fernandez | MD | Emergency Medicine | Clinical Professor and Executive Vice Chair of Academic Affairs and Faculty Development. | University of Florida, Gainesville, FL |
| Robert R. Gaiser | MD | Anesthesiology | Professor and Anesthesiology Residency Program Director; ABA Executive Director of Professional Affairs and member of the Board of Directors; Member of the ABA OSCE Committee. | Yale University, New Haven, CT |
| Michelle L, R. Gonzalez | PhD, CRNA, FAANA, CHSE-A | Nurse Anesthesia, Simulation Education | Director of the Nurse Anesthesia DNP Program, Clinical Associate Professor; AANA Simulation Subcommittee (founding member); International Nursing Association for Clinical Simulation and Learning (INACSL) Simulation Education Program (ISEP) Development Team. | University of Arkansas for Medical Sciences, College of Nursing, Little Rock, AR |
| Kenward B. Johnson | MD, MS | Anesthesiology | Professor of Anesthesiology, Adjunct Professor of Biomedical Engineering; ABA Examiner and member of the ABA OSCE Committee. | University of Utah, Salt Lake City, UT |
| Joan H. Kinniry | MSN, CRNP | Acute Care Nursing Practice | Advanced Practice Manager, Medical Intensive Care Unit | University of Pennsylvania, Philadelphia, PA |
| Edward S. Lee | MD | Hospital Medicine | Associate Clinical Professor of Medicine; Associate Program Director of the Internal Medicine Residency Training Program; Associate Chief of Education, Medical Services Greater Los Angeles VA | University of California, Los Angeles |
| Virginia (Chris) Muckler | DNP, CRNA, CHSE | Nurse Anesthesia | Associate Clinical Professor; Assistant Program Director of Nurse Anesthesia; National | Duke University, Raleigh, NC |

| | | | | |
|---|---|---|---|---|
| | | | League for Nursing Simulation Leader. | |
| David Murray | MD | Anesthesiology | Carol B and Jerome T Loeb Professor and Professor of Pediatric Anesthesiology [Emeritus], Former Director of the Howard and Joyce Wood Simulation Center [now retired] | Washington University, St. Louis, MO |
| Alexis Nicpon | MSN, RN, CPAN | Perioperative Nursing | Staff Nurse; Visiting Professor, Chamberlain College of Nursing. | Northwestern Medicine Central DuPage Hospital, Winfield, IL |
| Manuel Pardo | MD | Anesthesiology | Professor and Vice Chair for Education, Department of Anesthesia and Perioperative Care; ABA examiner. | University of California, San Francisco |

§ The 11 SMEs were nominated by a senior colleague and went through a rigorous vetting process. They were all board-certified in their clinical discipline (>5 years of practice experience), had a national reputation, a solid background in the education and evaluation of clinical trainees and/or practitioners, and had at least a general understanding of simulation methods. Following a modified Delphi process, using the custom web-based platform (VidARA™), SMEs first reviewed and commented on the content and nature of the draft simulation scenarios. After the scenarios were refined and pilot tested, the SMEs helped to generate the expected differential diagnoses (DDx) and clinical performance elements (CPEs) (i.e., medical/technical or behavioral actions expected) for each scenario. These DDx and CPEs were then ranked by the SMEs for importance, the results shared among all SMEs, and an online discussion occurred, which was followed by a second online ranking that yielded the final rating checklists.

#### SDC-4. Scenario Delivery Materials.

Complete scenario scripts and associated supporting materials to deliver them in a standardized manner are available from Drs. Weinger or Slagle upon written request.

For additional information, see (including Supplemental Digital Content):

1. McIvor, W. R., Banerjee, A., Boulet, J. R., Bekhuis, T., Tseytlin, E., Torsher, T., DeMaria Jr, S., Rask, J., Shotwell, M., Burden, A., Cooper, J., Gaba, D. M., Levine, A., Park, C., Sinz, E., Steadman, R., and Weinger, M. B.: A taxonomy of delivery and documentation deviations during delivery of high-fidelity simulations. *Simulation in Healthcare* 12(1): 1–8, Feb 2017. PMID: 28146449
2. Weinger, M. B., Banerjee, A., Burden, A., Boulet, J. R., Cooper, J., Steadman, R., McIvor, W. R., Shotwell, M., Slagle, J. M., DeMaria, S., Torsher, L., Sinz, L., Levine, A. I., Rask, J., Davis, F., Park, C., and Gaba, D. M.: Simulation-based assessment of the management of critical events by board-certified anesthesiologists. *Anesthesiology* 127(3): 475-89, Sep 2017. PMID: 28671903
3. Rask, J. P., Duran, H-T. [M], DeClercq, J., Andreae, M., Anders, S., Banerjee, A., Burden, A., Levine, A., Shotwell, M. S., Sinz, E., Torsher, L., Gaba, D. M., and Weinger, M. B.: “The prevalence, patterns and predictors of hazardous attitudes among anesthesiologists: An incompletely studied area of clinical decision safety.” *British Journal of Anaesthesia* 2023 Nov;131(5):e157-e160. Doi: 10.1016/j.bja.2023.08.012. PMID: 37741719.
4. Militello, L. G., Salwei, M. E., Reale, C., Sushereba, C., Slagle, J. M., Gaba, D., Weinger, M. B., Rask, J., Faiman, J., Andreae, M., Burden, A. R., and Anders, S.: Adapting cognitive interviewing techniques for use in a large sample study of high-risk healthcare events. *Journal of Cognitive Engineering and Decision Making* 2023 Dec;17(4):315-331. Doi: 10.1177/ 15553434231192283.

### SDC-5. End of Study Day Participant Survey Questions

#### A. Cognitive Interview eSurvey

| | |
|---|---|
| 1. Today's cases were relevant to my practice. | Strongly disagree / disagree / undecided / agree / strongly agree |
| 2. In my practice, I spend time after unusual clinical events reflecting (by myself or with others) on what might have been done differently. | Strongly disagree / disagree / undecided / agree / strongly agree |
| 3. The cognitive interviewers helped me to reflect upon the decisions I made during the simulated clinical events. | Strongly disagree / disagree / undecided / agree / strongly agree |
| 4. I enjoyed the cognitive interview process. | Strongly disagree / disagree / undecided / agree / strongly agree |
| 5. I felt psychologically safe enough to be candid during the cognitive interviews. | Strongly disagree / disagree / undecided / agree / strongly agree |
| 6. How did the guided self-reflection you did during the cognitive interviews affect your learning? | Free text |
| 7. What I learned as a result of the cognitive interviews may change my practice. | Strongly disagree / disagree / undecided / agree / strongly agree |
| 8. A cognitive interview like what you did today would be useful after actual critical events. | Strongly disagree / disagree / undecided / agree / strongly agree |
| 9. Do you have any other comments? | Free text |

#### B. Course Evaluation eSurvey

| | |
|---|---|
| 1. This experience met its stated objectives | Strongly disagree / disagree / undecided / agree / strongly agree |
| 2. Do you feel this program conveyed any commercial bias? | Yes/No |
| 3. The course content was relevant to my practice | Strongly disagree / disagree / undecided / agree / strongly agree |
| 4. The experience was non-threatening and conducive to learning. | Strongly disagree / disagree / undecided / agree / strongly agree |
| 5. The simulated clinical situations were realistic. | Strongly disagree / disagree / undecided / agree / strongly agree |
| 6. The program faculty and staff were effective in facilitating my learning. | Strongly disagree / disagree / undecided / agree / strongly agree |
| 7. I had an opportunity to reflect on my performance. | Strongly disagree / disagree / undecided / agree / strongly agree |
| 8. This was an effective use of my time. | Strongly disagree / disagree / undecided / agree / strongly agree |
| 9. The format was an effective way to learn this material. | Strongly disagree / disagree / undecided / agree / strongly agree |

| | |
|---|---|
| 10. I expect to make changes in my practice as a result of this program. | Strongly disagree / disagree / undecided / agree / strongly agree |
| 11. If you are going to make changes, what are they? | Free text |
| 12. Did you discern any commercial bias in this program? | Yes/No |
| 13. Overall, this program was a positive learning experience. | Strongly disagree / disagree / undecided / agree / strongly agree |

#### SDC-6. VidARA Video Management Software

VidARA (**Vid**eo **A**ssessment and **R**esearch **A**pplication) is an innovative video data management software application, designed for scientific, education and industry applications, that facilitates distributed comprehensive assessment and analysis of any type of videorecording. VidARA enables users to seamlessly download, organize, review, rate, and annotate videos all linked to the video timeline. The platform accommodates up to four video uploads, intelligently merged into a split-screen view for comparative analysis. VidARA allows user management, video data management (including linkage to external databases such as REDCap), secure assignment and management of remote raters, remote analyses and ratings, and export of reports including annotations and scores. Separate notes about training experience, videos, and raters can be created, aggregated and reviewed. Access of each type of user can be pre-specified based on their designated role (e.g., administrator, auditor, trainer, rater). A built-in notification system ensures timely task completion by sending email reminders to video auditors or raters about new assignments. VidARA data are stored in secure servers using state-of-the-art HIPAA and FERPA compliant processes. Overall, VidARA offers a powerful solution that simplifies video-based evaluation and data management.

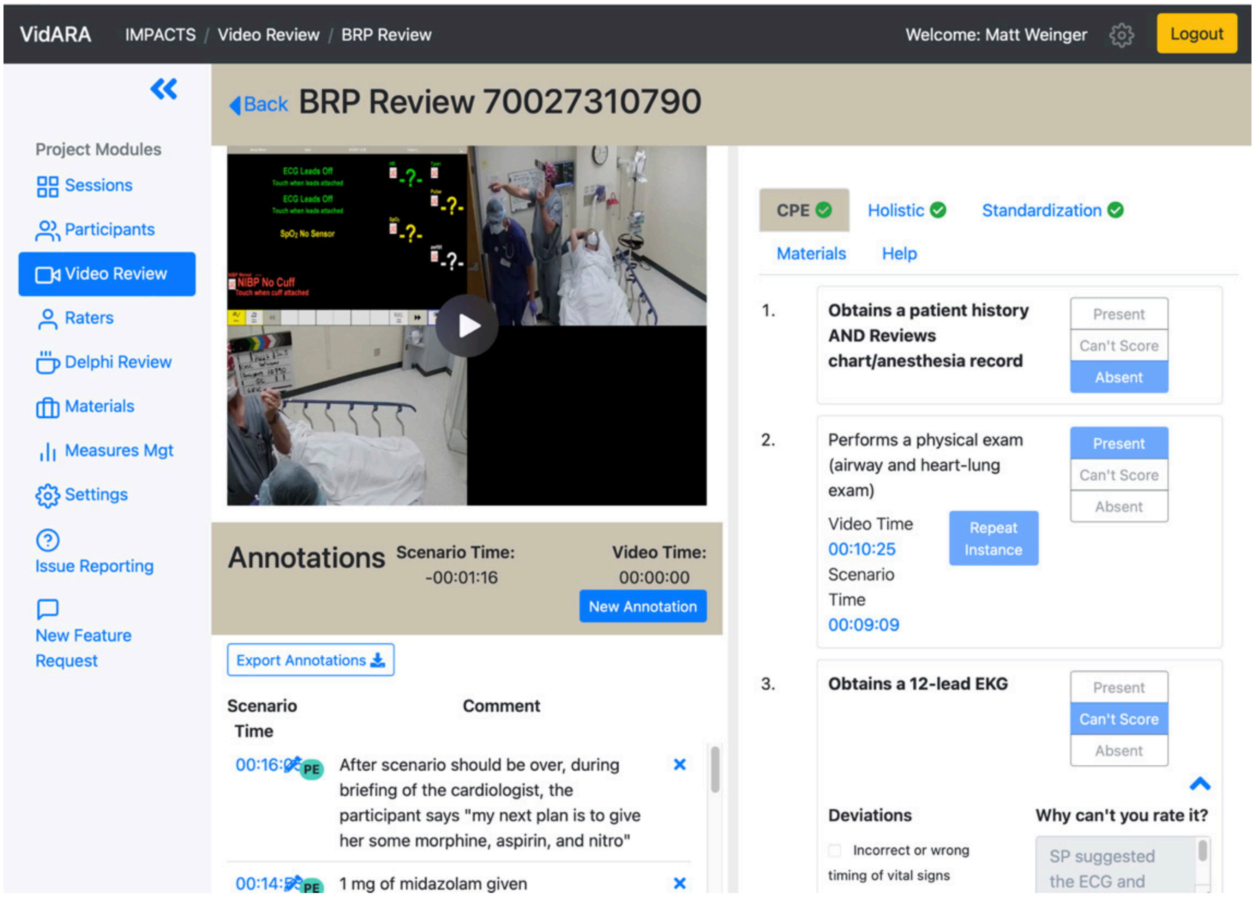


**Figure S2. ViDARA screen during CPE scoring of an actual study scenario**

#### SDC-7. Table S2. Clinical Performance Elements for Each Scenario

| # | SOB | Hypotension | Altered Mental Status | Anxiety |
|---|---|---|---|---|
| 1 | Obtains a patient history AND Reviews chart/anesthesia record | Obtains a patient history AND Reviews chart/anesthesia record | Asks anesthesiologist about patient history and possible precipitants | Obtains a patient history AND Reviews chart/anesthesia record |
| 2 | Performs a physical exam (airway, heart-lung, OR abdominal exams) | Performs a physical exam (airway, heart-lung, OR abdominal exams) | Orders additional labs (based on reasonable diagnoses) | Performs a physical exam (airway and heart-lung exam) |
| 3 | Escalates supplemental oxygen levels | Gives intravenous fluid bolus at least 500cc | Obtains a blood glucose level | Obtains a 12-lead EKG |
| 4 | Calls for clinician assistance [MD, RN, or other] | Administers a pressor (bolus) or starts an infusion | Conducts more extensive neurological exam | Recognizes and articulates myocardial ischemia |
| 5 | Gives intravenous fluid bolus of at least 500cc | Escalates supplemental oxygen levels | Attempts to dissuade/prevent confederate anesthesiologist from giving naloxone and/or flumazenil | Orders additional labs (based on reasonable diagnoses) |
| 6 | Administers a pressor (bolus) or starts an infusion | Obtains an ABG | Articulates need to broaden the differential diagnosis (beyond the common diagnoses in this situation) | Administers supplemental oxygen |
| 7 | Obtains a chest X-Ray | Obtains a 12-lead EKG | Administers sedative/hypnotic | Escalates supplemental oxygen levels |
| 8 | Obtains an ABG | Obtains a chest X-Ray | Considers either serotonin syndrome OR neuroleptic malignant syndrome in differential diagnosis. | Orders cardiac enzymes |
| 9 | Secures or is prepared to imminently secure the airway | Contacts urologist to discuss patient status and concerns | Orders administration of cyproheptadine (Periactin™) or chlorpromazine (Thorazine™) | Administers a pressor (bolus) |

| | | | | |
|---|---|---|---|---|
| 10 | Obtains a 12-lead EKG | POC Ultrasound to evaluate cardiac, lungs, abdomen | | Calls cardiology |
| 11 | POC Ultrasound to evaluate cardiac, lungs, abdomen | Calls for clinician assistance [MD, RN, or other] | | Administers additional pressors (bolus or infusion) |
| 12 | Escalates care by either calling for an ICU bed or contacting surgeon to discuss further management | Orders additional labs (based on reasonable diagnoses) | | Asks for POC ultrasound to evaluate cardiac function |
| 13 | Orders additional labs (based on reasonable diagnoses) | Gives additional volume | | |
| 14 | Escalates hemodynamic support | Secures or is prepared to imminently secure the airway | | |
| 15 | Contact Blood Bank | Escalates hemodynamic support | | |
| 16 | Reviews and broadens antibiotic coverage | Reviews and broadens antibiotic coverage | | |
| 17 | | Gives stress dose steroids | | |

#### SDC-8. Behaviorally Anchored Rating Scales (BARS) Criteria

*The descriptions (anchors) for the five BARS domains for poor, middling, and excellent performance*. Described observable behaviors for Poor, Average, and Excellent performance on the first two axes (domains) of the BARS instrument used in rating IMPACTS scenarios.

| Axes Elements | Poor | Average | Excellent |
|---|---|---|---|
| **Axis 1: Vigilant Observation** | Rarely pays attention to relevant data streams | More often than not pays attention to relevant data streams | Nearly always pays attention to all relevant data streams |
| **Vigilance:** Sustained attention (i.e., no serious lapses)<br>**Observation:** Attending to data streams<br>**Verification:** Estimating truth / falsehood of observation (e.g., vs. artifact or transient) | Rarely detects subtle but detectable cues | More often than not detects subtle detectable cues | Nearly always detects subtle detectable cues |
| | Rarely verifies observations by cross-checking or appropriate watchful waiting | More often than not verifies observations by cross-checking or watchful waiting | Nearly always verifies observations by cross-checking or watchful waiting |
| **Axis 2: Situation Awareness and Re-evaluation** | Rarely makes appropriate sense of relevant data | More often than not makes appropriate sense of relevant data | Nearly always makes appropriate sense of relevant data |
| **Situation Awareness – 3 levels** *(see Gaba et al Hum Factors 1995 and extensive work by Micah Endsley and others)*<br>Detecting anomalies and events<br>• Attributing "meaning" to observations<br>Assimilating many data streams/ pieces of information (including those not seemingly immediately relevant)<br>Anticipating and preparing for future events<br>• Predicting future states<br>Re-evaluation of the situation (also consistent with Klein's RPD)<br>• Changes in severity<br>• +/– Response to treatment<br>• New problems | Rarely grasps the bigger picture | More often than not grasps the bigger picture | Nearly always grasps of the bigger picture |
| | Rarely projects future possible problem | More often than not projects future possible problem evolution | Nearly always projects future possible problem evolution |
| | Rarely re-evaluates the situation (including about problem evolution or +/– response to Rx) | More often than not re-evaluates the situation (including about problem evolution or +/– response to Rx) | Nearly always re-evaluates the situation (including about problem evolution or +/– response to Rx) |
| **Axis 3: Diagnostic Processes** | Rarely recognizes abnormal observations as problems | More often than not recognizes abnormal observations as problems | Nearly always recognizes abnormal observations as problems |
| **Recognizing 'Problems':** "This observation suggests that there is a 'problem.'" *This is a special aspect of attribution of meaning. [as per Axis 2]*<br>**Retrieving "pre-compiled" set of likely potential causes**<br>**Generating abstract hypotheses of possible causes:** Diagnoses / Etiologies *[see also Gary Klein: Recognition Primed Decision Making (RPD); also Kahneman: Type I Decision Making]*<br>**Considering alternatives**<br>**Seeking disconfirming evidence about putative diagnoses; reasons using 1st principles** *[per Kahneman Type II and other models]* | Limited set of pre-compiled diagnostic possibilities for problems recognized | Moderate set of precompiled diagnostic possibilities for problems recognized | Rich set of pre-compiled diagnostic possibilities for problems recognized |
| | Rarely generates / considers additional hypotheses for problems recognized | More often than not generates / considers additional hypotheses for problems recognized | Nearly always generates / considers additional hypotheses for problems recognized |
| | Rarely tests possibilities or hypotheses against data for problems recognized | More often than not tests possibilities or hypotheses against data for problems recognized | Nearly always tests possibilities or hypotheses against data for problems recognized |
| **Axis 4: Action Processes** | Limited set of possible actions | Moderate set of possible actions | Rich set of possible actions |
| **Choose and plan actual set of actions (from universe of possible actions) based on combined consideration of:**<br>Efficacy of candidate action<br>Urgency of problem *[e.g., some actions only applicable to highly urgent problems]*<br>Precondition/constraints *[e.g., can't start epi infusion until it is assembled and ready to go]*<br>Reversibility *[e.g., adenosine vs. Ca++ channel blocker]*<br>Time needed to apply or to take effect *[e.g., IV morphine vs. oral morphine]*<br>Side effects and contraindications *[e.g., typically useful actions may be inapplicable]*<br>**Schedule and manage actions:**<br>Which first, which later<br>Managing resources including long-latency actions<br>Interleaving actions<br>Handling interruptions<br>Recognizing "Problems"<br>• Action implementation<br>Who is capable of doing the action?<br>• Are they present, willing, and able?<br>Resources for implementation of the action<br>• What resources are needed to do it? Ensuring current or future availability<br>• Prospective memory to implement when available<br>"Doing it" *[executing the action – successful or not]*<br>*{NOTE: failure to fully implement key actions physically is not measurable in this study}* | Rarely considers pros and cons of different actions | More often than not considers pros and cons of different actions | Nearly always balances pros and cons of different actions |
| | Set or sequence of actions is illogical | Set or sequence of actions is more often than not logical | Set or sequence of actions is nearly always logical |
| | Rarely plans and sequences relevant actions | More often than not plans and sequences relevant actions | Nearly always or sequences relevant actions |
| | Rarely maintains action sequences and prospective memory if challenged with interruptions | More often than not maintains action sequences and prospective memory if challenged with interruptions | Nearly always maintains action sequences and prospective memory if challenged with interruptions |
| | Rarely begins to implement reasonable temporizing actions for life-critical situations in a timely fashion | More often than not begins to implement reasonable temporizing actions for life-critical situations in a timely fashion | Nearly always begins to implement reasonable temporizing actions for life-critical situations in a timely fashion |
| **Axis 5: Communication** | Rarely listens carefully to speech of others in team | More often than not listens carefully to speech of others in team | Nearly always listens carefully to speech of others in team |
| **Hearing → Listening → Understanding**<br>Listening to others at all levels of hierarchy<br>Inviting input from others<br>**Thinking → Speaking → [to be heard, etc.]**<br>Asking vs. Requesting vs. Ordering<br>Clearly identifying target of speech (use names if possible)<br>Crafting speech carefully (choose words wisely when possible)<br>Assertiveness with respect<br>**Closed Loop Communication**<br>Verifying heard and understood by "read back"<br>Verifying actions done by verbal reports<br>**Shared Mental Model**<br>Thinking out loud when appropriate<br>Conducting periodic VERBAL "re-evaluation" of situation with team members | Often misunderstands what is said | Sometimes misunderstands what is said | Rarely misunderstands what is said |
| | Rarely clarifies unclear statements or requests FROM others | More often than not clarifies unclear statements or requests FROM others | Nearly always clarifies unclear statements or requests FROM others |
| | Rarely enacts closed loop communication | More often than not enacts closed loop communication | Nearly always enacts closed loop communication |
| | Rarely makes precise and clear statements or requests TO others | More often than not makes precise and clear statements or requests TO others | Nearly always makes precise and clear statements or requests TO others |
| | Rarely considers credible ideas expressed by others | Sometimes considers credible ideas expressed by others | Nearly always considers credible ideas expressed by others |
| | Rarely conveys own thoughts, situation, or plan to others (including verbal re-evaluation) | More often than not conveys own thoughts, situation, or plan to others (including verbal re-evaluation) | Nearly always conveys own thoughts, situation, or plan to others (including verbal re-evaluation) |

**Figure S3. Detailed enumeration of behaviors associated with poor-middling-excellent performance on the five BARS axes.**

BARS were scored on a 9-point scale from 1 (worst) to 9 (best). To reach that determination, raters first chose whether the observed performance was "poor" (1-3), "medium" (4-6), or "excellent" (ratings 7-9). After that initial determination, they decided if the performance was poor, medium or excellent relative to the initial determination. For example, a rater might view a performance as "excellent", but rate it as the lowest performance that qualifies as excellent, resulting in a score of 7.

#### SDC-9. Principal Component Analysis

We hypothesized that performance ratings would be affected by the clinical practice and event management experience of participants, adjusting for potential confounders of these relationships. To address this hypothesis, three sets of composite variables, representing: 1) general clinical experience, 2) crisis event management experience, and 3) experience with simulation, were separately created using principal component analysis of the survey-derived variables.

##### Table S3. Experience Domain and Covariates' Loading on the first two Principal Components

| Experience Domain | Question | Levels | Contribution to PC1 (%) | Contribution to PC2 (%) |
|---|---|---|---|---|
| **Clinical** | Are there any other characteristics at [work_site] that make the occurrence of unexpected acute events more common than in typical practice of elective surgery? | Yes, No | 4.3% | 0.2% |
| | Work at a Tertiary Care, Non-Academic Medical Center | Any, None | 14.4% | 0.3% |
| | Work at a Tertiary Care, Academic Medical Center | Any, None | 28.5% | 1.4% |
| | How often do you do you own cases | Frequently, Sometimes/ Never | 13.6% | 4.2% |
| | How often do you supervise providers in training | Frequently, Sometimes/ Never | 14.8% | 18.5% |
| | How often do you treat patients classified as ASA 3 or higher | Frequently, Sometimes/ Never | 11.4% | 3.0% |
| | Number of specialties/ subspecialties | Count | 1.9% | 23.3% |
| | Advanced fellowship training | Yes, No | 6.8% | 11.6% |
| | What is the average total number of hours you work each week? | Continuous | 4.2% | 22.5% |
| | How many hours each week do you spend doing clinical work (including hands on anesthesia care, direct supervision of nurse anesthetists, residents, or anesthesia assistants)? | Continuous | 0.2% | 15.1% |
| **Crisis** | Active involvement in hands-on crisis management drills | Any, None | 3.2% | 15.1% |

| | | | | |
|---|---|---|---|---|
| | How often did you encounter challenging situations in the past month? | Frequently, Sometimes/ Never | 34.5% | 1.0% |
| | How often did you encounter challenging situations in the past year? | Frequently, Sometimes/ Never | 32.7% | 0.5% |
| | How often do you work on patients who are acutely unstable | Frequently, Sometimes/ Never | 13.8% | 10.3% |
| | How often do you encounter emergencies like acute appendicitis or open fractures who are hemodynamically stable (2E or 3E) | Frequently, Sometimes/ Never | 15.7% | 4.2% |
| | Have you had any other formal training in clinical crisis management? | Yes, No | 0.1% | 35.3% |
| | Other relevant training to crisis management and teamwork? | Yes, No | 0.0% | 33.7% |
| **Simulation** | Have you completed a MOCA Simulation Course at an ASA endorsed center? | Yes, No | 4.5% | 26.2% |
| | Have you taken a hands-on anesthesia or critical care simulation course (e.g., Anesthesia-centric-Advanced Cardiac Life Support (A-ACLS)) at an academic meeting or simulation center? | Yes, No | 5.6% | 9.5% |
| | Have you taken any other type of simulation-based course? | Yes, No | 25.3% | 9.0% |
| | Simulation coursework which included teamwork and crisis management? | Yes, No | 39.7% | 7.0% |
| | Have you had any other formal training in team based clinical skills that involved active role playing? | Yes, No | 7.5% | 4.3% |
| | Have you worked with the ASA's SimStat screen-based simulation training modules? | Yes, No | 0.5% | 21.6% |
| | Training experience in simulation coursework? | Yes, No | 17.0% | 22.3% |

*Methods*: To reduce the dimensionality of more than 100 experience related variables while retaining features most relevant to the planned analyses, we used a semi-curated approach that combined expert guided variable selection with principal component analysis (PCA). An expert panel was convened to select the 7–10 potentially most informative variables for each of three different experience domains: routine clinical practice, crisis management, and simulation experience. The panel assessed both relevance to the study questions, as well as potential to differentiate participants.

This step prevented indiscriminate inclusion of all available variables and ensured that inputs to the PCA reflected domain specific content expertise.

PCA was then conducted separately within each domain to derive composite experience measures. All variables were centered and scaled prior to analysis. For each domain, the first two principal components were retained for subsequent modeling, balancing variance explained with interpretability. Biplots were used to examine loading patterns and identify the variables contributing most strongly to PC1 and PC2. This reduced >100 original potential covariates to six principal component scores (two per domain) while preserving interpretable structure in the resulting components.

#### SDC-10. Table S4. Rater Reliability Results

| Outcome | Anxiety | SOB | Hypotension | AMS | Overall |
|---|---|---|---|---|---|
| Intra-rater reliability | | | | | |
| CPE * | 0.867 | 0.925 | 0.85 | 0.818 | 0.858 |
| BARS ‡ | 0.849 | 0.604 | 0.815 | 0.558 | 0.759 |
| Medical/Technical Performance ‡ | 0.94 | 0.484 | 0.837 | 0.569 | 0.779 |
| Non-technical Performance ‡ | 0.902 | 0.58 | 0.931 | 0.484 | 0.790 |
| Overall Performance ‡ | 0.898 | 0.612 | 0.824 | 0.590 | 0.773 |
| Consultant Level Performance *† | 0.542 (0.699) | 0.545 (0.655) | 0.621 (0.662) | -0.077 (0.835) | 0.521 (0.735) |
| Inter-rater reliability | | | | | |
| CPE § | 0.847 | 0.869 | 0.860 | 0.776 | 0.848 |
| BARS § | 0.643 | 0.412 | 0.550 | 0.595 | 0.554 |
| Medical/Technical Performance § | 0.645 | 0.312 | 0.577 | 0.647 | 0.554 |
| Non-technical Performance § | 0.677 | 0.466 | 0.510 | 0.57 | 0.558 |
| Overall Performance § | 0.695 | 0.406 | 0.514 | 0.718 | 0.593 |
| Consultant Level Performance § † | 0.590 (0.00) | 0.400 (0.00) | 0.412 (0.00) | 0.626 (0.00) | 0.485 (0.00) |

Analysis method used:

* Cohen's kappa. Generally, values above 0.8 are considered 'excellent' and those above 0.60 are considered 'good' or 'substantial.'

† For the Consultant Level Performance (CLP) ratings, because kappa is sensitive to prevalence and marginal imbalance, especially of binary ratings (i.e., 78% of all CLP ratings were positive), Gwet's AC1 is shown (in parenthesis) as a complementary measure of reliability. With this approach, values above 0.60 are similarly considered 'good' or 'substantial.'

‡ Interclass correlation coefficient (ICC). Generally, values above 0.9 are considered excellent, those 0.75 to 0.90 are considered 'good', and those 0.50 to 0.75 are considered 'moderate'.

§ Krippendorff's alpha. Generally, values above 0.80 are considered 'good' or better.

#### SDC-11. Table S5. Summary of Performance Ratings by Scenario and Experience Cohort

| Performance by Scenario | N | AMS N = 102 | Anxiety N = 102 | Hypotension N = 102 | SOB N = 101 | P-value** |
|---|---|---|---|---|---|---|
| **CPEs (% completed)** | 407 | 57±8% (67, 56–78) | 62±5% (58, 50–67) | 56±7% (59, 53–65) | 58±8% (56, 50–63) | **0.001** |
| **BARS 1: Vigilance** | 407 | 6.3±1.6 (7, 5–7) | 5.8±1.6 (6, 5–7) | 5.8±1.9 (6, 4–7) | 6.1±1.6 (6, 5–7) | 0.093 |
| **BARS 2: Situation Awareness** | 406 | 5.8±2 (6, 4–7) | 5.8±1.7 (6, 5–7) | 5.4±2.2 (6, 4–7) | 5.7±1.8 (6, 4–7) | 0.372 |
| **BARS 3: Diagnostic Process** | 407 | 5.8±1.9 (6, 5–7) | 5.7±1.8 (6, 5–7) | 5.1±2 (5.5, 3–7) | 5.3±1.8 (6, 4–7) | **0.006** |
| **BARS 4: Action Processes** | 407 | 5.8±1.8 (6, 4–7) | 5.6±1.6 (6, 4–7) | 5.4±1.9 (6, 4–7) | 5.6±1.7 (6, 4–7) | 0.455 |
| **BARS 5: Communication** | 407 | 6.3±1.8 (7, 5–8) | 6.5±1.5 (7, 6–8) | 6.1±1.8 (6, 5–8) | 6.3±1.6 (7, 5–8) | 0.309 |
| **Medical/Technical Performance** | 407 | 5.9±1.9 (6, 4–7) | 5.6±1.7 (6, 4–7) | 5.4±2 (6, 3–7) | 5.6±1.7 (6, 4–7) | 0.202 |
| **Non-technical Performance** | 407 | 6±1.7 (6.5, 5–7) | 6±1.7 (6, 5–7) | 5.5±2 (6, 4–7) | 5.8±1.8 (6, 4–7) | 0.180 |
| **Overall Performance** | 406 | 5.9±1.8 (6, 5–7) | 5.8±1.6 (6, 5–7) | 5.5±2 (6, 4–7) | 5.6±1.8 (6, 4–7) | 0.281 |
| **Consultant Level Performance** | 407 | 81% (83) | 81% (83) | 72% (73) | 78% (79) | 0.251 |

| Performance by Experience Cohort | N | BCA N = 243 | Senior N = 76 | Junior N = 88 | Overall N = 407 | P-value** |
|---|---|---|---|---|---|---|
| **CPEs (% completed)** | 407 | 57±8% (57, 52–63) | 62±5% (61, 59–65) | 56±7% (54, 52–61) | 58±8% (57, 52–63) | **0.045** |
| **BARS 1: Vigilance** | 407 | 5.9±1.7 (6, 5–7) | 5.8±1.7 (6, 4–7) | 6.3±1.6 (7, 5–7.5) | 6±1.7 (6, 5–7) | 0.343 |
| **BARS 2: Situation Awareness** | 406 | 5.7±1.9 (6, 4–7) | 5.4±1.9 (5, 3.5–7) | 6.0±1.9 (6, 5–8) | 5.7±1.9 (6, 4–7) | 0.211 |
| **BARS 3: Diagnostic Process** | 407 | 5.4±1.9 (6, 4–7) | 5.3±1.9 (5, 4–7) | 6.0±1.9 (7, 5–7) | 5.5±1.9 (6, 4–7) | **0.049** |
| **BARS 4: Action Processes** | 407 | 5.6±1.7 (6, 4–7) | 5.3±1.8 (5, 4–7) | 6.0±1.7 (6, 5–7) | 5.6±1.7 (6, 4–7) | 0.079 |
| **BARS 5: Communication** | 407 | 6.2±1.7 (7, 5–7) | 6.1±1.7 (6, 5–8) | 6.7±1.5 (7, 6–8) | 6.3±1.7 (7, 5–8) | 0.212 |
| **Medical/Technical Performance** | 407 | 5.6±1.8 (6, 4–7) | 5.3±1.9 (5, 4–7) | 6.1±1.7 (7, 5–7) | 5.6±1.8 (6, 4–7) | 0.088 |

| | | | | | | |
|---|---|---|---|---|---|---|
| **Non-technical Performance** | 407 | 5.8±1.8 (6, 4–7) | 5.6±1.8 (6, 4–7) | 6.3±1.8 (7, 5–8) | 5.8±1.8 (6, 4–7)* | 0.078 |
| **Overall Performance** | 406 | 5.6±1.8 (6, 4–7) | 5.4±1.8 (5, 4–7) | 6.1±1.7 (7, 5–8) | 5.7±1.8 (6, 4–7) | 0.083 |
| **Consultant Level Performance** | 407 | 78% (190) | 72% (63) | 86% (65) | 78% (318) | 0.152 |

Except for CPEs and Consultant Level performance (% yes), all data are presented as average ± standard deviation (median, interquartile range).

* = The correlation between technical and nontechnical performance is very high: r = 0.925. In a linear mixed model with a random intercept for participant, nontechnical scores were significantly higher than technical scores by an average of 0.22 points (mean difference = 0.22, 95% CI 0.14 - 0.30, $p < 0.001$).

** CPE: Binomial generalized linear mixed-effects model; BARS and continuous holistic scores: linear mixed effects models; Consultant level performance: mixed effects logistic regression

#### SDC-12. Table S6. Summary of Holistic Score Regression Results

| Medical/Technical Performance | Level | Estimate | Statistic | 95% CI | P | Global P |
|---|---|---|---|---|---|---|
| Scenario | Anxiety | — | — | — | — | 0.205 |
| | Altered mental status | 0.24 | 1.02 | -0.22, 0.69 | 0.308 | |
| | Shortness of breath | -0.06 | -0.262 | -0.51, 0.39 | 0.794 | |
| | Hypotension | -0.25 | -1.11 | -0.71, 0.20 | 0.269 | |
| Experience | Senior | — | — | — | — | 0.185 |
| | BCA | -0.45 | -1.15 | -1.2, 0.32 | 0.252 | |
| | Junior | -0.64 | -1.81 | -1.3, 0.06 | 0.073 | |
| Gender | Male | — | — | — | — | |
| | Female | 0.12 | 0.524 | -0.34, 0.58 | 0.601 | |
| Clinical experience | Clinical experience (PC1) | 0.01 | 0.143 | -0.15, 0.17 | 0.887 | 0.983 |
| | Clinical experience (PC2) | 0.01 | 0.079 | -0.20, 0.22 | 0.937 | |
| Simulation experience | Simulation experience (PC1) | 0.03 | 0.361 | -0.15, 0.22 | 0.719 | 0.915 |
| | Simulation experience (PC2) | -0.03 | -0.231 | -0.26, 0.21 | 0.818 | |
| Crisis experience | Crisis experience (PC1) | 0.15 | 1.68 | -0.03, 0.32 | 0.097 | 0.116 |
| | Crisis experience (PC2) | 0.12 | 1.14 | -0.09, 0.34 | 0.257 | |
| **Non-technical Performance** | **Level** | **Estimate** | **Statistic** | **95% CI** | **P** | **Global P** |
| Scenario | Anxiety | — | — | — | — | 0.182 |
| | Altered mental status | 0.03 | 0.128 | -0.42, 0.48 | 0.898 | |
| | Shortness of breath | -0.19 | -0.809 | -0.64, 0.27 | 0.419 | |
| | Hypotension | -0.42 | -1.83 | -0.87, 0.03 | 0.068 | |
| Experience | Senior | — | — | — | — | 0.192 |
| | BCA | -0.51 | -1.36 | -1.3, 0.24 | 0.177 | |
| | Junior | -0.59 | -1.7 | -1.3, 0.10 | 0.092 | |
| Gender | Male | — | — | — | — | |
| | Female | 0.09 | 0.417 | -0.36, 0.55 | 0.677 | |
| Clinical experience | Clinical experience (PC1) | 0 | 0.014 | -0.16, 0.16 | 0.989 | 0.996 |
| | Clinical experience (PC2) | -0.01 | -0.085 | -0.22, 0.20 | 0.932 | |
| Simulation experience | Simulation experience (PC1) | 0.07 | 0.786 | -0.11, 0.25 | 0.434 | 0.725 |
| | Simulation experience (PC2) | -0.02 | -0.2 | -0.25, 0.21 | 0.842 | |
| Crisis experience | Crisis experience (PC1) | 0.16 | 1.8 | -0.02, 0.33 | 0.076 | 0.061 |
| | Crisis experience (PC2) | 0.16 | 1.46 | -0.06, 0.37 | 0.147 | |

| Overall Performance | Level | Estimate | Statistic | 95% CI | P | Global P |
|---|---|---|---|---|---|---|
| Scenario | Anxiety | — | — | — | — | 0.284 |
| | Altered mental status | 0.13 | 0.55 | -0.32, 0.57 | 0.583 | |
| | Shortness of breath | -0.11 | -0.474 | -0.56, 0.34 | 0.636 | |
| | Hypotension | -0.3 | -1.34 | -0.75, 0.14 | 0.183 | |
| Experience | Senior | — | — | — | — | 0.195 |
| | BCA | -0.47 | -1.22 | -1.2, 0.29 | 0.224 | |
| | Junior | -0.61 | -1.75 | -1.3, 0.08 | 0.083 | |
| Gender | Male | — | — | — | — | |
| | Female | 0.1 | 0.459 | -0.35, 0.56 | 0.647 | |
| Clinical experience | Clinical experience (PC1) | 0.02 | 0.23 | -0.14, 0.18 | 0.818 | 0.972 |
| | Clinical experience (PC2) | -0.01 | -0.11 | -0.22, 0.20 | 0.913 | |
| Simulation experience | Simulation experience (PC1) | 0.03 | 0.358 | -0.15, 0.21 | 0.721 | 0.937 |
| | Simulation experience (PC2) | 0 | 0.023 | -0.23, 0.23 | 0.981 | |
| Crisis experience | Crisis experience (PC1) | 0.16 | 1.88 | -0.01, 0.34 | 0.064 | **0.04** |
| | Crisis experience (PC2) | 0.18 | 1.65 | -0.04, .39 | 0.103 | |
| **Consultant Level Performance** | **Level** | **Odds Ratio** | **Statistic** | **95% CI** | **P** | **Global P** |
| Scenario | Anxiety | — | — | — | — | 0.242 |
| | Altered mental status | 1 | 0 | 0.48, 2.08 | >0.999 | |
| | Shortness of breath | 0.81 | -0.59 | 0.40, 1.65 | 0.556 | |
| | Hypotension | 0.54 | -1.73 | 0.27, 1.08 | 0.083 | |
| Experience | Senior | — | — | — | — | 0.298 |
| | BCA | 0.55 | -1.15 | 0.20, 1.53 | 0.249 | |
| | Junior | 0.5 | -1.49 | 0.20, 1.24 | 0.136 | |
| Gender | Male | — | — | — | — | |
| | Female | 1.13 | 0.39 | 0.62, 2.06 | 0.696 | |
| Clinical experience | Clinical experience (PC1) | 1.02 | 0.212 | 0.83, 1.26 | 0.832 | 0.978 |
| | Clinical experience (PC2) | 1 | -0.029 | 0.76, 1.30 | 0.977 | |
| Simulation experience | Simulation experience (PC1) | 1.11 | 0.914 | 0.88, 1.41 | 0.361 | 0.645 |
| | Simulation experience (PC2) | 1.04 | 0.24 | 0.78, 1.38 | 0.81 | |
| Crisis experience | Crisis experience (PC1) | 1.32 | 2.14 | 1.02, 1.71 | 0.032 | **0.015** |
| | Crisis experience (PC2) | 1.31 | 1.92 | 1.0, 1.72 | 0.054 | |

**SDC-13. Figure S4. Relationship of the percentage of CPEs performed and assigned overall global score for each scenario.**

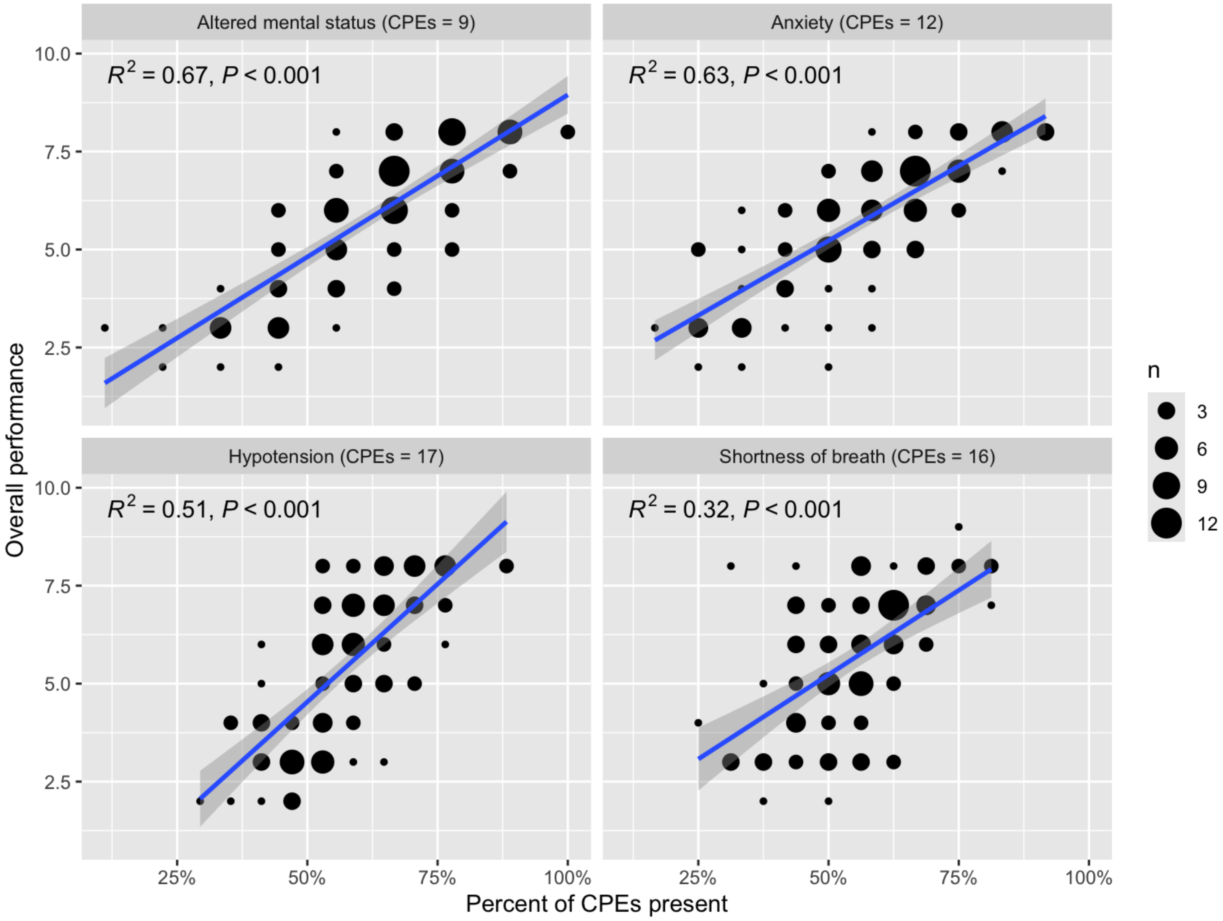


**Figure Legend.** For each of the four scenarios, this figure shows each participant's percentage of CPEs successful rated as performed vs. that participant's global score. In all cases, the more CPEs performed, the higher the global score with the relationship being the greatest for the *AMS* scenario (which had the fewest CPEs) and lowest for the *SOB* scenario. Spearman's rank correlation was additionally calculated as a descriptive measure of association. The proportion of CPEs marked as complete was strongly associated with overall performance ratings (Spearman's $\rho = 0.72$, $p < 0.001$).

#### SDC-14. Table S7. Clinical Performance Elements (CPEs) and Clinical Action Categories Associated with Higher Global Performance Scores - Overall and by Scenario

| Category of CPEs * | # CPEs | Percent of total CPEs | Average percent CPEs completed ˚ | # significant association ‡ | % significant association ‡ |
|---|---|---|---|---|---|
| **Combined** | | | | | |
| Initial evaluation | 6 | 11.1% | 88.4% | 0 | 0.0% |
| Initial diagnosis | 2 | 3.7% | 74.0% | 2 | 100.0% |
| Initial therapy | 6 | 11.1% | 87.4% | 4 | 66.7% |
| Seeks assistance | 5 | 9.3% | 46.3% | 3 | 60.0% |
| Diagnostic testing | 16 | 29.6% | 50.1% | 9 | 56.2% |
| Broaden DDx | 3 | 5.6% | 67.6% | 3 | 100.0% |
| Escalate therapy | 15 | 27.8% | 41.4% | 12 | 80.0% |
| Other | 1 | 1.9% | 49.0% | 0 | 0.0% |
| *Combined* | *54* | *100.0%* | *57.6%* | *33* | *61.1%* |
| **Shortness of breath** | | | | | |
| Initial evaluation | 2 | 12.5% | 98.5% | 0 | 0.0% |
| Initial diagnosis | 0 | | | | |
| Initial therapy | 3 | 18.8% | 81.5% | 2 | 66.7% |
| Seeks assistance | 2 | 12.5% | 23.8% | 1 | 50.0% |
| Diagnostic testing | 5 | 31.2% | 46.3% | 2 | 40.0% |
| Broaden DDx | 0 | | | | |
| Escalate therapy | 4 | 25.0% | 39.1% | 3 | 75.0% |
| Other | 0 | | | | |
| *Combined* | *16* | *100.0%* | *54.8%* | *8* | *50.0%* |
| **Hypotension** | | | | | |
| Initial evaluation | 1 | 5.9% | 99.0% | 0 | 0.0% |
| Initial diagnosis | 1 | 5.9% | 86.3% | 1 | 100.0% |
| Initial therapy | 2 | 11.8% | 91.7% | 1 | 50.0% |
| Seeks assistance | 2 | 11.8% | 55.4% | 1 | 50.0% |
| Diagnostic testing | 5 | 29.4% | 47.1% | 3 | 60.0% |
| Broaden DDx | 0 | | | | |
| Escalate therapy | 6 | 35.3% | 44.1% | 4 | 66.7% |
| Other | 0 | | | | |
| *Combined* | *17* | *100.0%* | *57.6%* | *10* | *58.8%* |

| | | | | | |
|---|---|---|---|---|---|
| **Anxiety** | | | | | |
| Initial evaluation | 2 | 16.7% | 67.6% | 0 | 0.0% |
| Initial diagnosis | 0 | | | | |
| Initial therapy | 1 | 8.3% | 96.1% | 1 | 100.0% |
| Seeks assistance | 1 | 8.3% | 72.5% | 1 | 100.0% |
| Diagnostic testing | 4 | 33.3% | 46.3% | 2 | 50.0% |
| Broaden DDx | 1 | 8.3% | 75.5% | 1 | 100.0% |
| Escalate therapy | 3 | 25.0% | 39.5% | 3 | 100.0% |
| Other | 0 | | | | |
| *Combined* | *12* | *100.0%* | *56.9%* | *8* | *66.7%* |
| **Altered mental status** | | | | | |
| Initial evaluation | 1 | 11.1% | 99.0% | 0 | 0.0% |
| Initial diagnosis | 1 | 11.1% | 61.8% | 1 | 100.0% |
| Initial therapy | 0 | | | | |
| Seeks assistance | 0 | | | | |
| Diagnostic testing | 2 | 22.2% | 74.5% | 2 | 100.0% |
| Broaden DDx | 2 | 22.2% | 63.7% | 2 | 100.0% |
| Escalate therapy | 2 | 22.2% | 40.7% | 2 | 100.0% |
| Other | 1 | 11.1% | 49.0% | 0 | 0.0% |
| *Combined* | *9* | *100.0%* | *63.1%* | *7* | *77.8%* |

***** The individual CPEs in each category and the significance of their association with higher global scores can be found in **Supplemental Digital Content, Table XS**.

° Calculated as the weighted average based on the number of individual CPEs in each group.

‡ The statistical criteria for significance was p<0.05 but about one third of associations were at the p≤0.001 level and about 80% were at the p≤0.02 level.

† This was the CPE “Attempts to dissuade/prevent confederate anesthesiologist from giving naloxone and/or flumazenil” in the AMS scenario which did not fit into the other broad categories and did not show a statistically significant association.
